# Machine learning-derived Alzheimer’s disease dimensions: neuroanatomical, cognitive, clinical, functional, and genetic risk heterogeneity signatures

**DOI:** 10.64898/2026.09.02.26361668

**Authors:** Hoang Nam Le, Saba Ishrat, Bahram Yaghooti, Taslim Murad, Ram Sapkota, Deepa S. Thakuri, Dean F. Wong, Carlos Cruchaga, Alzheimer’s Disease Neuroimaging Initiative, Ganesh B. Chand

**Author notes:** Corresponding authors: Hoang Nam Le PhD and Ganesh B. Chand PhD.

## Abstract

Alzheimer’s disease (AD) is characterized by substantial clinical and biological heterogeneity, with diverse neuroanatomical, cognitive, clinical, functional, and genetic risk profiles that are poorly captured by current diagnostic frameworks. Identifying reproducible disease dimensions is critical for understanding divergent pathophysiological mechanisms and designing future stratified therapeutic interventions. Herein, we applied a semi-supervised machine learning clustering framework and robust reproducibility validation strategies to structural MRI data from 1,757 participants, comprising 1,108 cognitively normal controls (CN) and 649 patients with mild cognitive impairment (MCI) or dementia due to AD. We identified two MRI-based neuroanatomical dimensions of AD: Dimension 1 (*n* = 393; prevalence ∼ 61%) and Dimension 2 (*n* = 256; prevalence ∼ 39%). Dimension 1 exhibited widespread cortical and subcortical atrophy, including involvement of the hippocampus, amygdala, parahippocampal, temporal, frontal, and occipital regions, while Dimension 2 showed relatively preserved brain patterns. The two dimensions were not statistically different in demographic characteristics, including age, sex, race, and education (all p > 0.05). Compared to Dimension 2, Dimension 1 had more severe cognitive and clinical impairment, greater functional impairment, elevated *APOE ε4* carrier burden, and higher polygenic risk for AD (p < 0.05). Analysis of individual-level summary neuroanatomical signature expression scores, quantifying each subject’s continuous position along two disease dimensions, revealed that Dimension 1 signature expression correlated strongly with cognitive impairment, clinical severity, functional impairment, and genetic risk, while Dimensional 2 signature expression showed relatively weaker associations. These results reveal that AD encompasses at least two biologically distinct dimensions identifiable from structural MRI profiles and their distinct associations with cognitive, clinical, functional, and genetic profiles. These findings may aid in AD patient stratification for designing targeted therapeutic approaches in the future.

## 1 INTRODUCTION

Dementia affects more than 57 million people worldwide, with Alzheimer’s disease (AD) being its most common cause, and remains a major cause of disability and mortality among older adults.^1^ According to the 2026 Alzheimer’s Disease Facts and Figures report, an estimated 7.4 million people aged 65 and older are living with AD dementia in the United States, and AD ranks as the fifth-leading cause of death among Americans aged 65 and older.^2^ Despite decades of research, there is no cure for AD; disease-modifying treatments have proven elusive,^3^ and a growing body of evidence suggests that biological heterogeneity may contribute to the complexities of diagnosis, prognosis, and therapeutic development.^4–6^ Current diagnostic criteria and treatment paradigms treat AD as a largely homogeneous entity, yet AD patients can present with markedly different patterns of neurodegeneration, cognitive decline trajectories, and other profiles, potentially suggesting the presence of biologically distinct disease dimensions. Such disease heterogeneity may have important implications for future clinical trial design. By aggregating biologically distinct patient subpopulations under a single diagnostic label, trials risk diluting treatment effects and obscuring dimension-specific responses, potentially contributing to variability in treatment responses and clinical trial outcomes.^7^ Moreover, even within identified dimensions, patients vary continuously in the degree to which they express a given disease pattern, suggesting that both categorical and continuous representations of heterogeneity dimensions are crucial.

Among imaging biomarkers, structural magnetic resonance imaging (MRI) is particularly well-suited for capturing AD heterogeneity, as it quantifies atrophy across all gray matter, white matter, and cerebrospinal fluid (CSF)/ventricular regions, providing a holistic macroscopic view of neurodegeneration.^8–10^ In contrast, other imaging approaches such as diffusion tensor imaging (DTI) are primarily focused on white matter microstructure,^11^ whereas positron emission tomography (PET) molecular imaging, while capable of measuring protein-specific pathology (for example, amyloid-beta and tau), is constrained by partial volume effects, off-target binding, and its sensitivity to only one tracer-specific protein at a time,^12^ missing the multifactorial contributions to AD heterogeneity. Structural MRI thus offers an aggregated perspective on disease variation and its associations with cognitive, clinical, functional, and genetic profiles. We therefore employed structural MRI to characterize neuroanatomical heterogeneity in AD.

To characterize AD heterogeneity using data-driven approaches, several neuroimaging-based studies have applied unsupervised and semi-supervised machine learning (ML) to CSF, PET, and structural MRI data, among others. Dong et al. identified distinct AD dimensions defined by differential cortical and hippocampal atrophy patterns using non-negative matrix factorization applied to structural MRI.^13^ Young et al. employed a data-driven event-based model to reconstruct individualized disease progression sequences from biomarker data, revealing substantial inter-patient variability in the ordering of pathological events.^14^ Zhang et al. applied a Bayesian model to identify overlapping latent factors representing distinct atrophy patterns in structural MRI.^15^ Vogel et al. used unsupervised clustering of tau-PET data to delineate spatially distinct tau spreading dimensions associated with different clinical outcomes and genetic risk profiles.^16^ Qiu et al. applied principal component analysis and model-based clustering to neuropsychological test data to identify typical and atypical cognitive profiles in probable AD, with atypicality associated with distinct genetic, neuropathological, and clinical progression trajectories.^17^ Together, these studies, among others, demonstrate that AD patients can be stratified into distinct heterogeneity dimensions. However, many existing methods rely on unsupervised algorithms that do not incorporate available diagnostic labels (both controls and patients). As a result, patients are stratified based on similarity metrics rather than true disease-related deviations from controls, and the resulting dimensions can be confounded by demographic differences, limiting their ability to capture disease-relevant biological heterogeneity. Furthermore, robust semi-supervised ML application and validation of AD heterogeneity across large, independent, and demographically diverse cohorts remains limited.

Semi-supervised ML offers a principled alternative by incorporating known case-control labels to guide the clustering solution. Rather than grouping patients by arbitrary distance metrics in feature space, semi-supervised ML approaches anchor cluster boundaries to what biologically differentiates patients from controls, thereby increasing the likelihood that identified dimensions capture disease-relevant variation. The heterogeneity through discriminative analysis (HYDRA) method implements this principle by simultaneously learning *k* support vector machine hyperplanes, each separating cognitively normal controls (CN) from a distinct patient dimension, with cluster membership determined by the maximum signed margin distance.^18^ The HYDRA framework, integrated with robust reproducibility strategies, has been proposed for identifying the optimal number of disease dimensions,^19^ and this integrated approach has been successfully applied to various psychiatric disorders, such as schizophrenia^19^, psychosis-spectrum,^20^ autism,^21^ and first-episode psychosis.^22^ However, such integrated HYDRA and robust reproducibility validation approaches remain to be systematically evaluated in AD for identifying optimal disease heterogeneity dimensions in large and diverse cohorts.

A complementary limitation of previous approaches is that they reduce each patient to a discrete cluster membership, obscuring the continuous variation in disease expression that exists within and across dimensions.^16^ In reality, individual patients likely differ not just in which dimension they belong to, but in the degree to which their neuroanatomy expresses each disease-dimensional pattern.^23,24^ Recent work in psychiatric neuroimaging has demonstrated that quantifying individual-level summary expression of heterogeneity signatures as continuous scores can reveal clinically meaningful variation beyond what dimensional membership alone conveys.^20^ Importantly, this individual-level summary perspective enables the investigation of relationships between neuroanatomical heterogeneity and various measures, such as cognition, clinical severity, functional impairment, and genetic risk, at the individual level rather than the dimensional level, providing a more personalized characterization of biological heterogeneity signatures in disease. Prior studies have not combined such robust reproducibility methods with validation across independent cohorts and investigated the identification of AD dimensions by quantifying individual-level expression of neuroanatomical signatures. Consequently, it remains unknown whether MRI-derived reproducible neuroanatomical patterns correspond to clinically and genetically meaningful dimensions of disease heterogeneity. Addressing these gaps may improve biological stratification and facilitate more precise approaches in the AD field.

In this study, we applied HYDRA integrated with robust reproducibility methods to a combined cohort drawn from the Alzheimer’s Disease Neuroimaging Initiative (ADNI)^25^ and Washington University’s Knight Alzheimer Disease Research Center (Knight ADRC),^26^ representing one of the largest semi-supervised ML and multimodal heterogeneity analyses in AD to date. We first identified reproducible neuroanatomical dimensions of AD and investigated their relationships with cognitive, clinical, functional, and genetic measures. The disease dimensions are defined based on their deviations from normative brain structure, facilitating the identification of neurobiologically meaningful patterns of disease variation. To validate the robustness of the discovered AD dimensions, we conducted multiple reproducibility analyses, including repeated cross-validation, permutation-based null hypothesis testing, and split-sample testing. We then quantified individual-level expression of the resulting neuroanatomical heterogeneity signatures in AD patients and examined their associations with cognitive, functional, clinical severity, and genetic risk factors. We hypothesized that our integrated approach would identify stable patterns of disease heterogeneity relative to CN and that the identified neuroanatomical heterogeneity signatures would be distinctly associated with cognitive, clinical, functional, and genetic risk characteristics.

## 2 MATERIALS AND METHODS

### 2.1 Study Sample

T1-weighted structural MRI datasets from the Knight ADRC (*n* = 1090) and ADNI (*n* = 667) were selected for this study. The sample included 1108 CN and 649 patients at various stages of disease: mild cognitive impairment (MCI, *n* = 6), early MCI (EMCI, *n* = 172), late MCI (LMCI, *n* = 152), or AD dementia (*n* = 319). Participants with a baseline T1 MRI scan and available baseline mini-mental state examination (MMSE) scores were included in the study. IRB approvals were obtained at each participating institution in accordance with the Declaration of Helsinki, and written informed consent was obtained from all participants or their authorized representatives prior to study enrollment. **Table 1** summarizes key demographic details of the studied population, including the race and ethnicity of participants in each cohort. Detailed inclusion and exclusion criteria are provided in a prior study^26^ and **Supplementary Parts A-B**.

**Table 1.** Demographic characteristics of the study sample stratified by cohorts.

|  | **Combined** | **ADNI** | **Knight ADRC** |
| --- | --- | --- | --- |
| *n* | 1757 | 667 | 1090 |
| Age, mean ± SD | 71.31 ± 8.24 | 72.83 ± 7.34 | 70.37 ± 8.62 |
| Female, *n* (%) | 914 (52.0%) | 308 (46.2%) | 606 (55.6%) |
| Years of education, mean ± SD | 16.14 ± 2.58 | 16.29 ± 2.61 | 16.04 ± 2.56 |
| **Diagnosis** |  |  |  |
| · CN, *n* (%) | 1108 (63.0%) | 187 (28.0%) | 921 (84.5%) |
| · EMCI, *n* (%) | 172 (9.8%) | 172 (25.8%) | 0 (0.0%) |
| · LMCI, *n* (%) | 152 (8.6%) | 152 (22.8%) | 0 (0.0%) |
| · MCI, *n* (%) | 6 (0.3%) | 6 (0.9%) | 0 (0.0%) |
| · AD Dementia, *n* (%) | 319 (18.1%) | 150 (22.5%) | 169 (15.5%) |
| **Race** |  |  |  |
| · American Indian/Alaskan, *n* (%) | 1 (0.1%) | 1 (0.1%) | 0 (0.0%) |
| · Asian, *n* (%) | 21 (1.2%) | 14 (2.1%) | 7 (0.6%) |
| · Black, *n* (%) | 179 (10.2%) | 27 (4.0%) | 152 (13.9%) |
| · Hawaiian/Other Pacific Islander, *n* (%) | 2 (0.1%) | 2 (0.3%) | 0 (0.0%) |
| · More than one, *n* (%) | 7 (0.4%) | 7 (1.0%) | 0 (0.0%) |
| · Unknown, *n* (%) | 1 (0.1%) | 1 (0.1%) | 0 (0.0%) |
| · White, *n* (%) | 1546 (88.0%) | 615 (92.2%) | 931 (85.4%) |
| **Ethnicity** |  |  |  |
| · Hispanic/Latino, *n* (%) | 33 (1.9%) | 29 (4.3%) | 4 (0.4%) |
| · Not Hispanic/Latino, *n* (%) | 1679 (95.6%) | 636 (95.4%) | 1043 (95.7%) |
| · Unknown, *n* (%) | 45 (2.6%) | 2 (0.3%) | 43 (4.0%) |

ADNI, Alzheimer's Disease Neuroimaging Initiative; Knight ADRC, Charles F. and Joanne Knight Alzheimer Disease Research Center; CN, cognitively normal; EMCI, early mild cognitive impairment; LMCI, late mild cognitive impairment; MCI, mild cognitive impairment; AD, Alzheimer's disease.

### 2.2 Image Preprocessing and Harmonization

T1-weighted structural MR images were segmented into 145 anatomical regions of interest (ROIs) using the multi-atlas region segmentation using ensembles of registration algorithms and parameters with locally optimal atlas selection (MUSE),^27^ a consensus labeling framework that employs a spatially adaptive label fusion strategy across multiple registered brain atlases. Image preprocessing steps are detailed in **Supplementary Part C**. These 145 regions cover the whole brain, including cortical and subcortical gray and white matter areas and CSF/ventricular regions. This whole-brain coverage is critical for unbiased discovery of holistic heterogeneous atrophy patterns. MUSE was chosen for segmentation because it outperforms traditional segmentation methods in multi-site settings.^28^ To account for site-related variation, scanner differences, and demographic confounders, harmonized ROI volumetric data were generated using ComBat-GAM^29,30^. This ComBat-GAM step adjusted the brain measures for acquisition site, cohort, age, sex, and total intracranial volume for each participant, with age modeled as a nonlinear covariate.

### 2.3 Study Variables

The primary features used for identifying AD neuroanatomical heterogeneity were the 145 regional brain volumetric measures described above. Age at MRI acquisition, biological sex, years of education, self-reported race, and ethnicity were recorded. Global cognitive function and clinical severity were assessed using the MMSE closest to the MRI scanning date,^31^ and the Clinical Dementia Rating (CDR) scale with the CDR sum of boxes (CDR-SB).^32^ CDR domains, covering memory, orientation, judgment and problem solving, community affairs, home and hobbies, and personal care, were also examined to study the domain-specific clinical severity profiles. Functional impairment was assessed using the Functional Activities Questionnaire (FAQ).^33^ For genetic variables, *apolipoprotein E (APOE)* genotype was encoded as *ε4* carrier status: non-carrier, heterozygous carrier (one *ε4*), or homozygous carrier (both *ε4*), and AD polygenic risk scores (PRS) were calculated separately.^34,35^ Additional clinical variables, including laboratory measures, history of cardiovascular disease, and other metabolic risk factors, were also examined (details in **Supplementary Part D-E**). Several variables were only partially available across cohorts or restricted to a single cohort due to differences in data collection protocols; a complete summary of missing data by cohort is provided in **Supplementary Table S1**.

### 2.4 Reproducible and Optimal Neuroanatomical Dimensions of AD

Semi-supervised clustering was performed using HYDRA,^18^ which simultaneously learns *k* discriminative hyperplanes separating CN from *k* patient dimensions. Each patient is assigned to an AD dimension corresponding to the hyperplane with the greatest signed margin, defined as the signed distance between the patient’s feature vector and the separating hyperplane. Controls contribute to the optimization of all hyperplanes but are not assigned to any AD dimension. A range of possible clustering solutions was investigated, specifically *k* = 2 to 8. The regularization parameter *C* = 0.25 was selected based on prior applications in neurological and psychiatric disorders. Semi-supervised clustering was applied with 10-fold cross-validation across 50 independent runs, and the optimization stability was quantified using the adjusted Rand index (ARI; 0 = chance, 1 = perfect reproducibility across sample resolutions). To confirm that identified dimensions reflect genuine biological structure rather than algorithmic artifacts, we conducted robust reproducibility analyses integrated with HYDRA following a previous study:^19^ (1) the full real dataset; (2) split-sample validation; (3) a balanced real-data subsampling, in which a balanced training dataset consisting of randomly sampled CN and patients was used; and (4) a simulated null condition generated by randomly permuting case-control labels. The optimal number of patient dimensions ‘*k’* was selected based on the highest and most stable mean ARI, with significance assessed against the permutation-derived null distribution. The HYDRA algorithm and its implementation are provided in **Supplementary Part F**, and details of each reproducibility experiment are provided in the **Supplementary Part G**.

### 2.5 Summary Neuroanatomical Heterogeneity Signature Expression of AD

Patient dimensions were identified by learning *k* discriminative hyperplanes, each of which separates controls from one patient dimension in the neuroanatomical feature space. Each hyperplane is defined by a weight vector *w_k_* and a bias term *b_k_*, estimated during training via a semi-supervised support vector machine formulation. As demonstrated in prior studies,^20,22,36^ the geometric structure of the clustering hyperplanes is a powerful tool for the quantification of the degree to which an individual expresses a disease-related neuroanatomical pattern (details in **Supplementary Part F**).

### 2.6 Statistical Analysis

Between-group differences in age, years of education, body mass index (BMI), CDR-SB, CDR domain scores, MMSE, FAQ, PRS, and other available lab measurements were compared using Mann-Whitney U tests. Categorical measures, such as sex, race, history of cardiometabolic and cardiovascular disease, history of smoking, diabetes, and *APOE ε4* allele frequency were compared using chi-square tests. Spearman’s correlation analysis was performed between neuroanatomical heterogeneity signature expressions and cognitive, clinical, functional, and genetic measures.

Regional atrophy profiles of disease dimensions were characterized by computing Cohen’s d effect size for each of the 145 ROIs relative to CN, with ROI-level significance assessed by Welch’s t-tests. Benjamini-Hochberg false discovery rate (FDR) correction (q < 0.05) was applied to account for multiple comparisons across the 145 ROIs. The reproducibility of the identified dimensions was quantified using the ARI across cross-validation folds, with significance assessed against a permutation-based null distribution. All statistical analyses were performed in Python (version 3.12) using the SciPy, statsmodels, and scikit-learn libraries.^37–39^

## 3 RESULTS

### 3.1 Two Optimal and Most Stable Neuroanatomical Dimensions of AD Revealed

We identified *k* = 2 as the optimal number of dimensions, achieving the highest mean ARI of 0.51 across 50 repetitions of 10-fold cross-validation (**Figure 1**). This solution significantly exceeded the null distribution derived from a randomly permuted label dataset (p < 0.001) and remained stable across split-sample (ARI = 0.29 ± 0.05 for each half) and balanced case-control experiments (ARI = 0.21 ± 0.05). Clustering stability ARIs decreased with increasing *k* (*k* = 3: 0.31; *k* = 5: 0.21; *k* = 8: 0.15), showing that additional dimensions did not improve clustering stability and that the two-dimensional solution provides the most reproducible representation of AD neuroanatomical heterogeneity in this large dataset. The two dimensions comprised 393 patients in Dimension 1 (prevalence ∼ 61%) and 256 patients in Dimension 2 (prevalence ∼ 39%).

**Figure 1.**
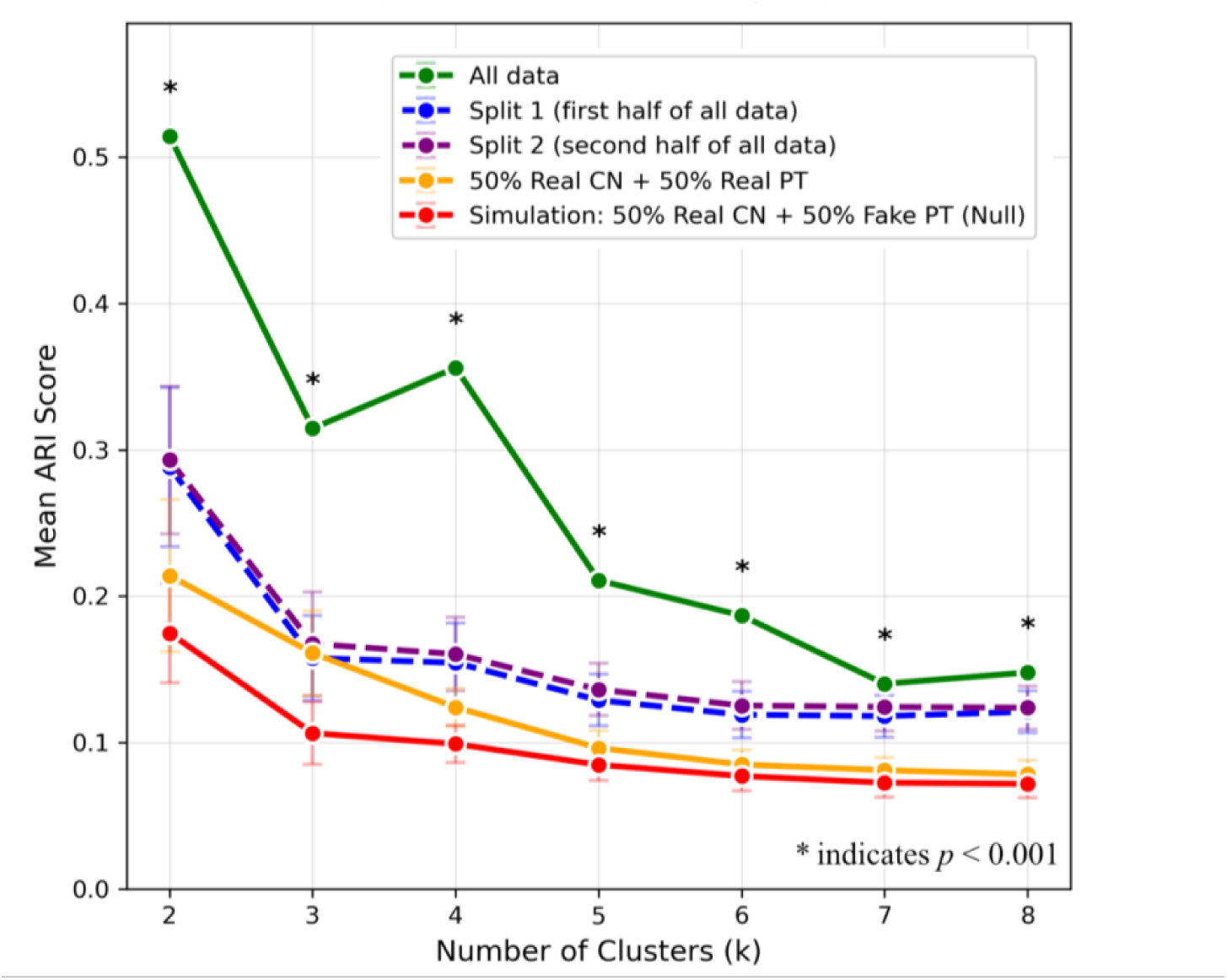
Reproducibility of semi-supervised clustering solutions across multiple validation experiments. Mean adjusted Rand index (ARI) across clustering solutions (*k* = 2 to 8) under five experimental conditions: the full dataset (green), split-sample validation (blue and purple), balanced case-control subsampling (orange), and the permutation-based null condition with randomly assigned patient labels (red). The two-dimensional solution (*k* = 2) achieved the highest reproducibility (mean ARI = 0.51) and significantly outperformed the null distribution (p < 0.001). Reproducibility decreased progressively with increasing numbers of dimensions, supporting *k* = 2 as the optimal solution. Error bars represent standard deviations across repeated runs. * indicates p < 0.001 versus the permutation-derived null distribution.

### 3.2 AD Dimensions Exhibited Distinct Neuroanatomical Patterns

The neuroanatomical characteristics of the two AD dimensions were examined by comparing regional brain volumes with CN. We computed Cohen’s d effect size between each AD dimension and CN across 145 gray matter, white matter, and CSF/ventricle regions (**Figure 2**). The two dimensions differed markedly in the spatial distribution and directionality of brain volumetric differences. Dimension 1 showed large positive effect sizes spanning nearly the entire brain, with the strongest differences involving the hippocampus, temporal and frontal cortices, and basal ganglia, reflecting diffuse, severe volumetric loss relative to CN. Dimension 2 showed a more spatially restricted and mixed profile: atrophy was evident in medial temporal and posterior regions, while several ventricular and periventricular structures showed larger volumes than CN.

**Figure 2.**
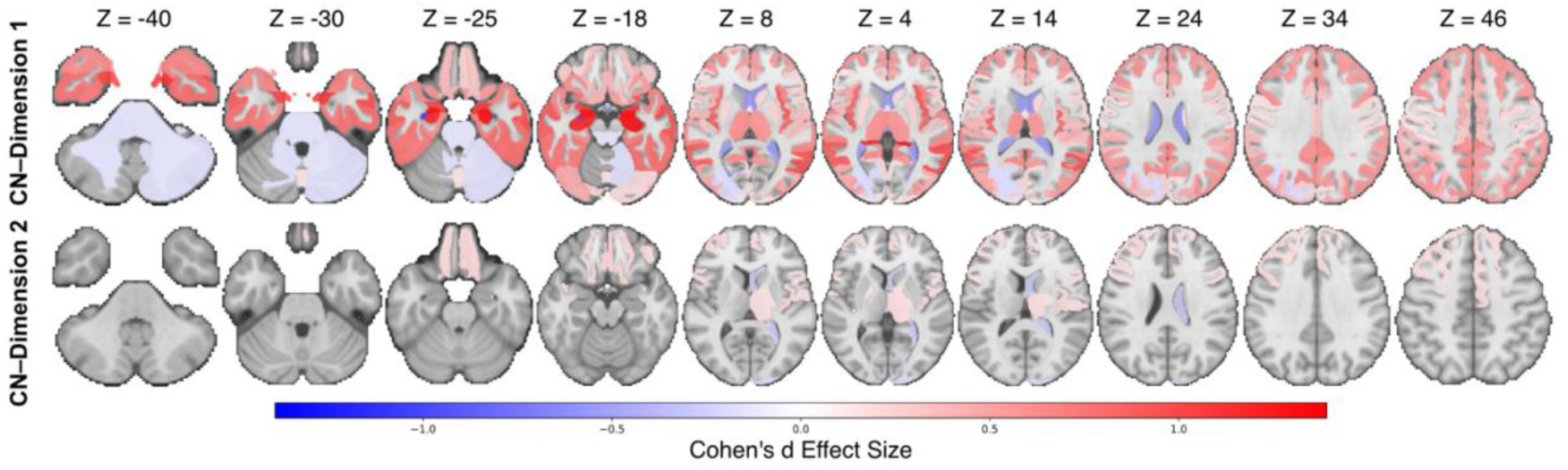
Regional neuroanatomical patterns of AD Dimensions 1 and 2 relative to cognitively normal controls (CN). Regional Cohen’s d effect size maps comparing Dimension 1 (*n* = 393) and Dimension 2 (*n* = 256) with CN (*n* = 1,108) across 145 brain regions. Dimension 1 exhibited widespread cortical and subcortical volume reductions, particularly involving temporal, medial temporal, frontal, and basal ganglia regions, together with ventricular enlargement. Dimension 2 showed relatively preserved brain structure, with more localized volume reductions and enlargement of ventricular/CSF regions. Positive values (red) indicate regional volume reduction relative to CN, whereas negative values (blue) indicate regional volume increase. Only regions with p < 0.05 are displayed, and color intensity represents Cohen’s *d* effect size. Slice positions are shown in MNI *z*-coordinates.

At the dimensional level, the neuroanatomical characteristics of Dimension 2 did not survive FDR correction across 145 ROIs, reflecting the smaller and more variable effect sizes in this dimension (**Supplementary Figure S1**). Sex-stratified analyses, however, revealed that significant regions emerged after correction when males and females were examined separately (**Supplementary Figure S2**). In Dimension 2, males showed robust ventricular enlargement surviving FDR correction, whereas no regions survived correction in females. In Dimension 1, the widespread atrophy pattern was broadly reproduced in both biological sexes, with males showing more periventricular volumetric changes compared to females, suggesting some sex-specific differences in neuroanatomical patterns observed across dimensions.

Among a total of 1,757 participants, 64 controls had non-zero baseline CDR-SB scores and 2 patients had zero baseline CDR-SB, reflecting discrepancies in the timing of score collection and diagnosis label assignment. Sensitivity analyses excluding these participants yielded neuroanatomical heterogeneity profiles (**Supplementary Part H**; **Supplementary Figure S3**) consistent with those from the full sample (**Figure 2**).

### 3.3 AD Neuroanatomical Dimensions Showed Divergent Cognitive, Clinical, Functional, and Genetic Risk Profiles

Patients in Dimension 1 demonstrated significantly greater global cognitive impairment, clinical severity, and functional impairment than those in Dimension 2 (**Table 2**, MMSE: mean (μ) = 24.7 vs. 27.3; CDR-SB: μ = 3.32 vs. 1.86; FAQ: μ = 9.14 vs. 3.34; all p < 0.001). These identified disease dimensions were not confounded by demographic differences: Age did not differ significantly (p = 1.000) between Dimension 1 (72.98 ± 7.91 years) and Dimension 2 (72.78 ± 7.27 years); sex distribution was similar, with Dimension 1 comprising 45.2% females and Dimension 2 comprising 41.8% females (χ^2^ = 0.590, p = 1.000); and years of education were also comparable between the two dimensions (15.79 ± 2.77 vs. 16.05 ± 2.80; p = 0.631).

**Table 2.**
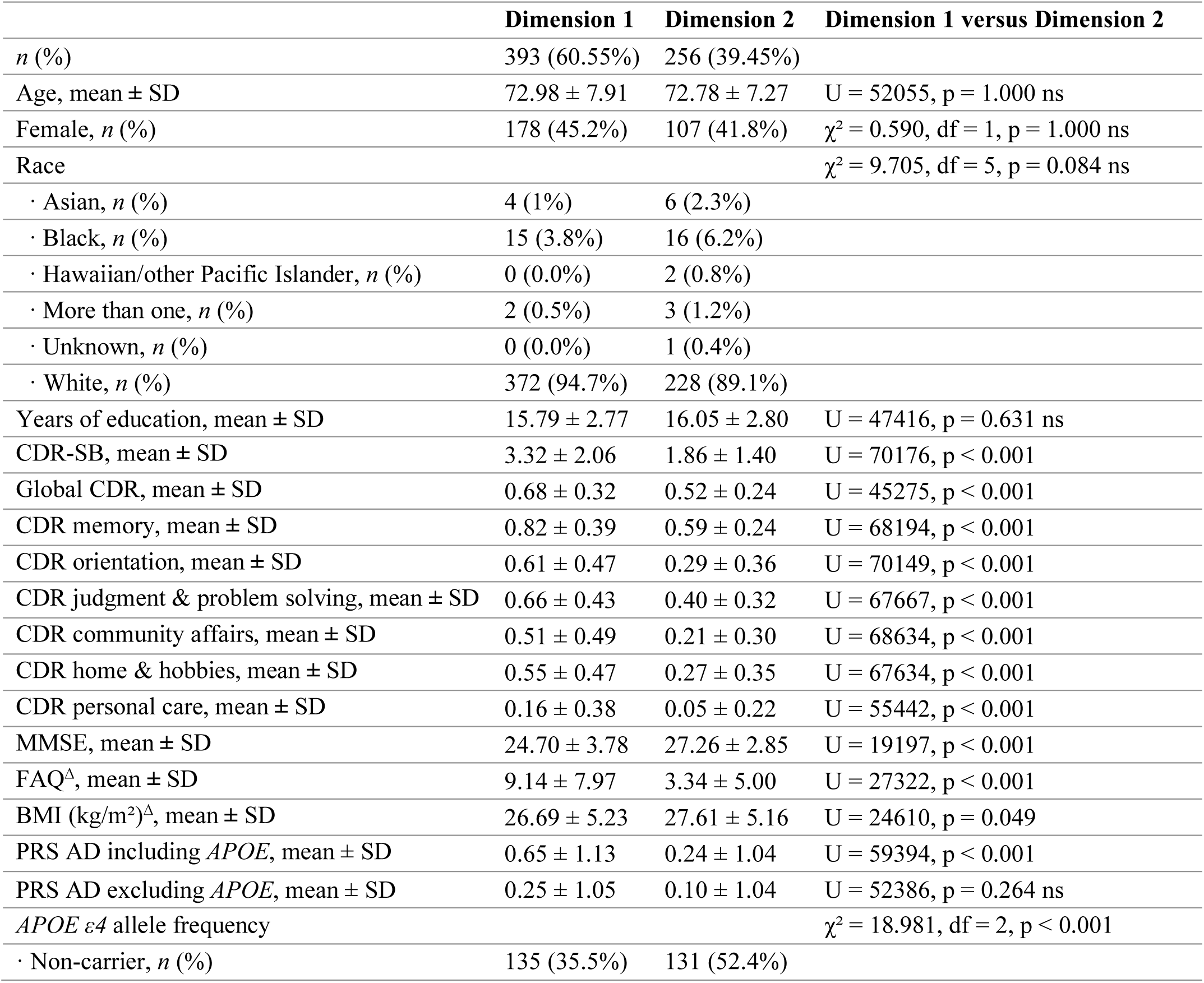
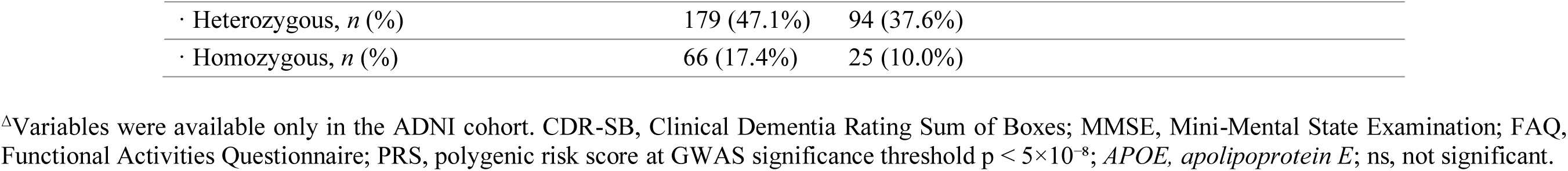
Comparisons between two AD neuroanatomical dimensions in demographic, cognitive, clinical, functional, and genetic characteristics.

|  | **Dimension 1** | **Dimension 2** | **Dimension 1 versus Dimension 2** |
| --- | --- | --- | --- |
| *n* (%) | 393 (60.55%) | 256 (39.45%) |  |
| Age, mean ± SD | 72.98 ± 7.91 | 72.78 ± 7.27 | U = 52055, p = 1.000 ns |
| Female, *n* (%) | 178 (45.2%) | 107 (41.8%) | $\chi^2$ = 0.590, df = 1, p = 1.000 ns |
| Race | | | $\chi^2$ = 9.705, df = 5, p = 0.084 ns |
| · Asian, *n* (%) | 4 (1%) | 6 (2.3%) |  |
| · Black, *n* (%) | 15 (3.8%) | 16 (6.2%) |  |
| · Hawaiian/other Pacific Islander, *n* (%) | 0 (0.0%) | 2 (0.8%) |  |
| · More than one, *n* (%) | 2 (0.5%) | 3 (1.2%) |  |
| · Unknown, *n* (%) | 0 (0.0%) | 1 (0.4%) |  |
| · White, *n* (%) | 372 (94.7%) | 228 (89.1%) |  |
| Years of education, mean ± SD | 15.79 ± 2.77 | 16.05 ± 2.80 | U = 47416, p = 0.631 ns |
| CDR-SB, mean ± SD | 3.32 ± 2.06 | 1.86 ± 1.40 | U = 70176, p < 0.001 |
| Global CDR, mean ± SD | 0.68 ± 0.32 | 0.52 ± 0.24 | U = 45275, p < 0.001 |
| CDR memory, mean ± SD | 0.82 ± 0.39 | 0.59 ± 0.24 | U = 68194, p < 0.001 |
| CDR orientation, mean ± SD | 0.61 ± 0.47 | 0.29 ± 0.36 | U = 70149, p < 0.001 |
| CDR judgment & problem solving, mean ± SD | 0.66 ± 0.43 | 0.40 ± 0.32 | U = 67667, p < 0.001 |
| CDR community affairs, mean ± SD | 0.51 ± 0.49 | 0.21 ± 0.30 | U = 68634, p < 0.001 |
| CDR home & hobbies, mean ± SD | 0.55 ± 0.47 | 0.27 ± 0.35 | U = 67634, p < 0.001 |
| CDR personal care, mean ± SD | 0.16 ± 0.38 | 0.05 ± 0.22 | U = 55442, p < 0.001 |
| MMSE, mean ± SD | 24.70 ± 3.78 | 27.26 ± 2.85 | U = 19197, p < 0.001 |
| FAQ<sup>A</sup>, mean ± SD | 9.14 ± 7.97 | 3.34 ± 5.00 | U = 27322, p < 0.001 |
| BMI (kg/m<sup>2</sup>)<sup>A</sup>, mean ± SD | 26.69 ± 5.23 | 27.61 ± 5.16 | U = 24610, p = 0.049 |
| PRS AD including *APOE*, mean ± SD | 0.65 ± 1.13 | 0.24 ± 1.04 | U = 59394, p < 0.001 |
| PRS AD excluding *APOE*, mean ± SD | 0.25 ± 1.05 | 0.10 ± 1.04 | U = 52386, p = 0.264 ns |
| *APOE* ε4 allele frequency | | | $\chi^2$ = 18.981, df = 2, p < 0.001 |
| · Non-carrier, *n* (%) | 135 (35.5%) | 131 (52.4%) |  |

|  |  |  |
| --- | --- | --- |
| · Heterozygous, *n* (%) | 179 (47.1%) | 94 (37.6%) |
| · Homozygous, *n* (%) | 66 (17.4%) | 25 (10.0%) |

<sup>Δ</sup>Variables were available only in the ADNI cohort. CDR-SB, Clinical Dementia Rating Sum of Boxes; MMSE, Mini-Mental State Examination; FAQ, Functional Activities Questionnaire; PRS, polygenic risk score at GWAS significance threshold $p < 5 \times 10^{-8}$ ; *APOE*, *apolipoprotein E*; ns, not significant.

Biological sex-stratified sensitivity analyses are presented in **Supplementary Table S2**. Specifically, within Dimension 1, male patients experienced a relatively greater burden in the CDR domains of home and hobbies (μ = 0.58 vs 0.51), and personal care (μ = 0.22 vs 0.08), while in Dimension 2, male patients showed greater impairment than females in CDR community affairs (μ = 0.23 vs 0.17). In both dimensions, males had more years of education than females. Within each dimension, females showed significantly higher lab cholesterol levels and lower lab creatinine as well as lab serum uric acid levels. A higher proportion of females than males in Dimension 1 had a history of endocrine-metabolic disease (52.3%).

Across the six CDR subdomains, Dimension 1 showed a pattern of greater impairment across daily cognitive and functional capacities (**Table 2**). Dimension 1 showed significantly higher impairment than Dimension 2 across all six domains (p < 0.001 each), including memory (μ = 0.82 vs. 0.59), orientation (μ = 0.61 vs. 0.29), judgment and problem solving (μ = 0.66 vs. 0.40), home and hobbies (μ = 0.55 vs. 0.27), community affairs (μ = 0.51 vs. 0.21), and personal care (μ = 0.16 vs. 0.05). No significant differences were observed in cardiometabolic markers and histories of cardiovascular and metabolic comorbidity between dimensions. However, individuals in Dimension 2 showed relatively higher triglyceride levels, although the difference was not statistically significant (**Supplementary Table S3**, μ =142.3 vs. 126.5 mg/dL, p = 0.083).

Dimension 1 had significantly higher AD PRS (PRS at the Genome-wide association studies (GWAS) significance threshold, p < 5×10⁻⁸) than Dimension 2 when *APOE* was included in the calculation (**Table 2**, μ = 0.65 vs. 0.24; p < 0.001), whereas no significant difference was observed when *APOE* was excluded from the PRS calculation. Among participants of European ancestry (*n* = 590), Dimension 1 had a significantly higher PRS than Dimension 2 when *APOE* was included in the score (0.67 ± 1.14 vs. 0.24 ± 1.07; p < 0.001), while APOE-excluded scores showed nonsignificant trend differences in the same direction (0.27 ± 1.07 vs. 0.13 ± 1.05; p = 0.117). AD dimensional profiles of PRS calculated at other GWAS significance thresholds (p < 5×10⁻⁵, 0.05, 0.5, and 1) are presented in **Supplementary Table S4**. Dimension 1 showed a significantly higher *APOE ε4* burden than Dimension 2 (χ^2^ = 18.981, p < 0.001), with 64.5% of Dimension 1 participants carrying at least one *APOE ε4* allele compared to 47.6% in Dimension 2. The enrichment was driven by both heterozygous (47.1% vs. 37.6%) and homozygous (17.4% vs. 10.0%) carriers, indicating a relatively higher association of Dimension 1 with *APOE ε4*-related genetic risk.

### 3.4 Neuroanatomical Dimensional Signature Expressions Correlated Distinctly with Cognitive, Functional, Clinical and Genetic Measures at Individual Level

At the individual level, Dimension 1 signature expression was associated with cognitive and functional impairment across the patient samples, as shown in **Figure 3**. Greater Dimension 1 signature expression was associated with lower MMSE (Spearman’s correlation (ρ) = −0.453, p < 0.001), greater functional impairment on the FAQ (ρ = 0.482, p < 0.001), and higher dementia severity on CDR-SB (ρ = 0.447, p < 0.001). In contrast, Dimension 2 signature expression showed a positive association with MMSE (ρ = 0.147, p < 0.001) and a weak negative association with CDR-SB (ρ = −0.110, p < 0.01), but no significant association with FAQ. Among laboratory biomarkers, greater Signature 1 expression was associated with lower serum uric acid (ρ = −0.103, p < 0.05) and lower triglyceride levels (ρ = −0.119, p < 0.05). Greater Signature 1 expression was also associated with lower BMI (ρ = −0.136, p < 0.01). For genetic markers, Dimension 1 signature expression was positively associated with *APOE*-inclusive AD PRS (ρ = 0.201, p < 0.001), while Dimension 2 signature expression showed a weak negative association with *APOE*-inclusive AD PRS (ρ = −0.087, p < 0.05). When the *APOE* region was excluded from the PRS calculations (**Supplementary Table S5**), Dimension 1 signature expression showed a nonsignificant positive association with AD PRS (ρ = 0.075, p = 0.058), while Dimension 2 signature expression showed no association with AD PRS (ρ = −0.014, p = 0.725).

**Figure 3.**
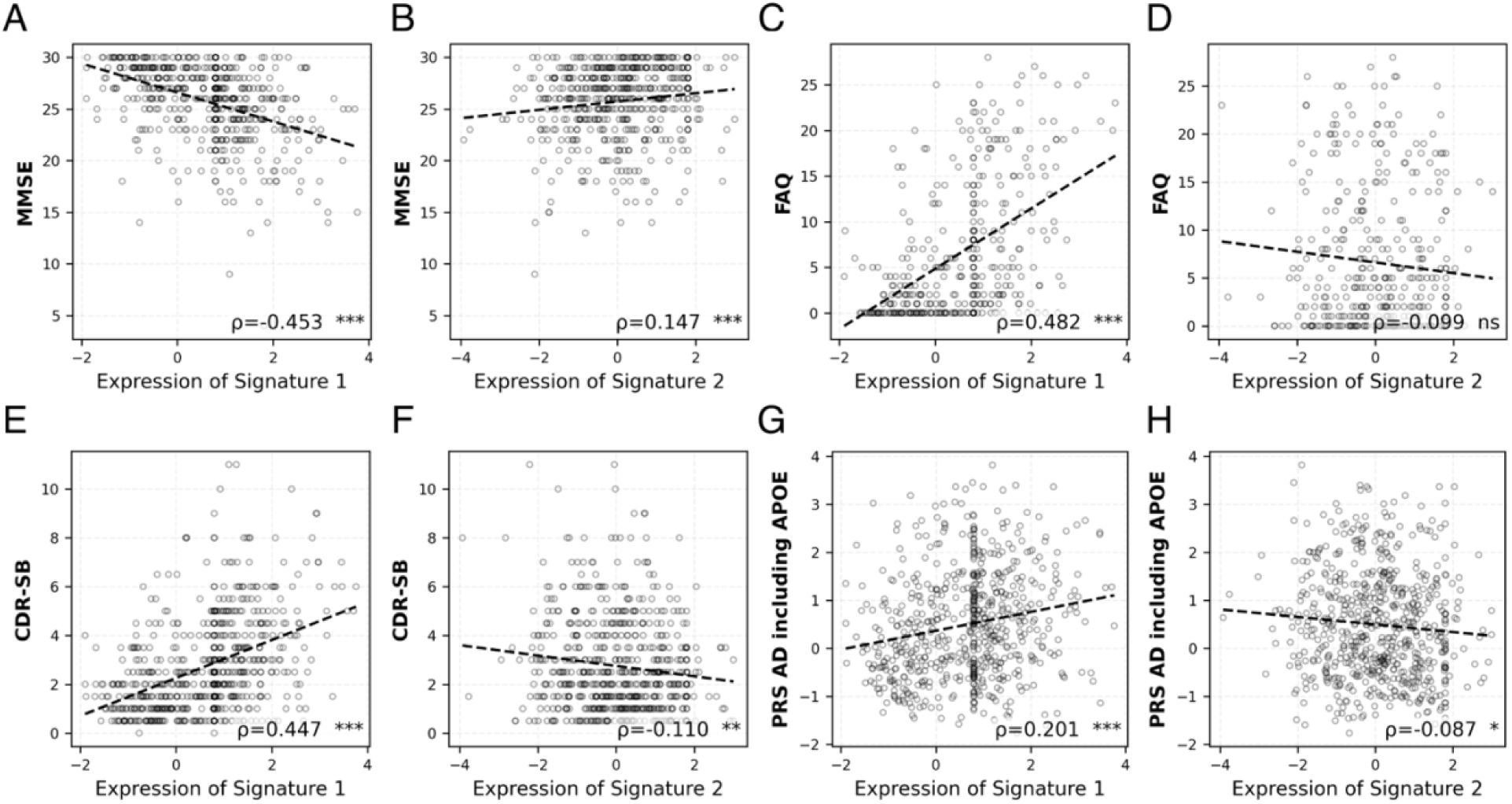
Scatter correlation plots between individual-level dimensional neuroanatomical signature expressions and cognitive (MMSE, A-B), functional (FAQ, C-D), clinical severity (CDR-SB, E-F), and genetic (PRS AD, G-H) measures in AD patients. Dimension 1 signature expression shows strong associations negatively with MMSE (Spearman’s ρ = −0.453, p < 0.001) and positively with FAQ (ρ = 0.482, p < 0.001), CDR-SB (ρ = 0.447, p < 0.001), and PRS for AD (ρ = 0.201, p < 0.001). Dimension 2 signature expression shows weak or non-significant positive associations with MMSE (ρ = 0.147, p < 0.001) and negative associations with PRS for AD (ρ = −0.087, p < 0.05). Each point represents one individual; dashed lines show ordinary least squares regression fits. Spearman’s correlation ρ and statistical significance are reported in the bottom-right corner of each panel. ns = not significant; * p < 0.05; ** p < 0.01; *** p < 0.001.

## 4 DISCUSSION

The present study identified two stable and reproducible neuroanatomical dimensions of AD heterogeneity in a large multi-cohort sample using semi-supervised machine learning and rigorous cross-validation approaches. The neuroanatomical profile of Dimension 1 was characterized by widespread volumetric reductions, including cortical and medial temporal atrophy. In contrast, Dimension 2 demonstrated relatively preserved brain structure. This study further demonstrated that two neuroanatomical dimensions exhibit distinct cognitive, clinical, functional, and genetic profiles, both at the dimensional level and individual level. The individual-level dimensional heterogeneity signature expressions and these distinct associations extended the characterization of AD heterogeneity beyond categorical dimensional assignment.

These two dimensions may reflect distinct neurobiological processes in AD: an atrophy-predominant pattern (Dimension 1) and a relatively preserved pattern (Dimension 2). This distinction is broadly consistent with prior data-driven heterogeneity studies of AD. Ferreira et al. identified typical, limbic-predominant, hippocampal-sparing, and minimal atrophy dimensions across neuropathological and neuroimaging studies, supporting the view that AD comprises multiple biologically distinct patterns of neurodegeneration rather than a single homogeneous disease process.^40^ Similarly, Dong et al. identified four neuroanatomical patterns in prodromal AD, including one with largely normal profiles and the slowest clinical progression, paralleling Dimension 2, and another with a classical AD profile and the fastest progression, paralleling Dimension 1.^13^ Consistent with this latter pattern, another work described a limbic-predominant tau spreading dimension in amyloid-positive individuals that maps closely onto the pattern observed in Dimension 1.^16^ Together, these findings support the interpretation that the widespread cortical and medial temporal involvement in Dimension 1 reflects a more severe neurodegenerative phenotype, contributing to its greater cognitive, clinical, and functional impairment, while the relative structural preservation in Dimension 2 points toward a distinct, and comparatively less severe, underlying process.

The identification of a relatively preserved neuroanatomical Dimension 2 is notable. Prior studies have reported hippocampal-sparing dimensions in which cognitive impairment is not accompanied by the expected pattern of structural atrophy,^41^ and which may reflect alternative or additional pathophysiological processes, including white matter disease,^42,43^ synaptic dysfunction or neuroinflammation,^44–46^ that can contribute to brain network dysfunction.^47,48^ The relative preservation of brain structure observed in Dimension 2 despite measurable cognitive, clinical, and functional impairment suggests that multiple neurobiological pathways can contribute to cognitive decline processes in AD. Consistent with this interpretation, Poulakis et al. identified multiple longitudinal atrophy trajectories in amyloid-positive AD and demonstrated that some cross-sectional atrophy patterns may reflect biologically distinct disease pathways rather than simply different stages of disease progression.^49^ In our approach, we quantified individual-level AD heterogeneity signature expression scores, which capture the extent to which each patient expresses each disease dimension. These findings indicate that the identified dimensions are not explained solely by disease severity. Rather, patients are grouped according to the direction of their neuroanatomical deviations from CN, while the signature scores quantify the degree of expression within each dimension. Consequently, patients with similar neuroanatomical patterns but different levels of disease severity can belong to the same dimension. In contrast to purely unsupervised clustering approaches, our approach incorporated the control group versus patient group membership while retaining the ability to identify neuroanatomically distinct patient dimensions. Furthermore, the use of two independent cohorts with complementary demographic and clinical characteristics provides additional support for the reproducibility and generalizability of the identified AD dimensions.

The divergence in cognitive and functional profiles between dimensions further supports the clinical relevance of the identified neuroanatomical patterns. Dimension 1 patients demonstrated significantly worse performance across all cognitive and functional measures, including MMSE, Global CDR, CDR-SB, FAQ, and all six CDR domains. The convergence of greater brain atrophy, worse cognitive performance, greater clinical severity, and higher functional impairment suggests that Dimension 1 represents a more severe neurodegenerative phenotype, whereas Dimension 2 represents a distinct clinical manifestation of disease. Our findings also suggested that vascular and metabolic factors may contribute less to the observed heterogeneity. These observed differences in cognitive and functional impairment suggest that clinical diagnosis alone may not fully capture underlying disease biology and support biologically informed stratification approaches in AD research and therapeutic development.^4^

The differential *APOE ε4* burden between dimensions is consistent with the established role of *APOE ε4* in accelerating neurodegeneration. The enrichment of *APOE ε4* heterozygous and homozygous carriers in Dimension 1 aligns with its widespread cortical and medial temporal atrophy patterns, whereas Dimension 2 showed a higher proportion of *APOE ε4* non-carriers despite meeting clinical criteria for MCI or AD. *APOE ε4* remains the strongest common genetic risk factor for AD and is thought to increase disease risk through multiple mechanisms, including neurodegeneration.^50^ Beyond *APOE* genotype, the PRS analyses provide additional insight into the genetic basis of the identified neuroanatomical heterogeneity signatures. At both the dimensional and individual levels, the association between Dimension 1 and AD polygenic risk appeared to be largely driven by *APOE*-related genetic risk, consistent with the established role of *APOE* as the strongest common genetic risk factor for AD.^51^ Dimension 2 showed only a weak association with *APOE*-inclusive PRS and no significant association with *APOE*-excluded PRS, despite the presence of cognitive and functional impairment.

Sex-stratified sensitivity analyses revealed some clinical severity differences within the dimensions. These differences were more apparent in atrophy-predominant Dimension 1, suggesting that the influence of sex on clinical expression may be shaped, in part, by underlying neuroanatomical phenotype rather than AD diagnosis alone. Although prior studies suggest that females may experience faster rates of cognitive decline than males,^52,53^ our findings indicate that the overall degree of cognitive and functional severity is comparable between males and females, consistent with literature reporting inconclusive evidence for sex and gender differences in cognitive decline trajectories.^54^

In contrast to the notable between-dimension differences observed across neuroanatomical, cognitive, clinical, functional, and genetic profiles, the cardiovascular risk factors, BMI, routine laboratory measures, and demographic characteristics were broadly comparable between Dimension 1 and Dimension 2. This suggests that these general health and lifestyle factors and demographics may not substantially distinguish between the two disease dimensions in the present sample and are unlikely to account for the divergent patterns of brain atrophy and cognitive impairment observed between them. Nonetheless, sex-stratified examination within each dimension revealed subtle differences in metabolic and renal biomarkers. Within both dimensions, females showed higher cholesterol levels than males, while males exhibited higher creatinine and uric acid levels than females. These findings suggest that the two identified dimensions are unlikely to be explained solely by sex-related differences in these metabolic markers. Thus, our results capture distinct information about AD heterogeneity rather than simply reflecting sex composition differences between the groups.

A key contribution of the present work is the quantification of neuroanatomical signature expression at the individual level, enabling a continuous characterization of neuroanatomical disease expression rather than a categorical dimensional assignment alone. Although this study robustly characterized neuroanatomical heterogeneity dimensions in AD and demonstrated their distinct cognitive, clinical, functional, and genetic associations, several limitations should be acknowledged. First, the study population was predominantly white (European ancestry), so future studies should evaluate the generalizability of the findings across more diverse racial and ancestral groups. Additionally, the current analysis is cross-sectional, precluding direct assessment of longitudinal stability, disease progression, or conversion trajectories. Future studies incorporating longitudinal neuroimaging, other biomarkers, and clinical follow-up will be important to determine how these heterogeneity dimensions evolve over time. Second, risk factors such as exercise-related physical activity^55^ and waist circumference and waist-to-hip ratio,^56^ which may provide more sensitive measures of adiposity-related risk than BMI alone, were not available in either cohort, limiting assessment of potential adiposity-related heterogeneity. Third, although multiple clustering solutions were evaluated, the present study focused on the most reproducible two-dimensional solution. It remains possible that larger datasets may support the identification of additional biologically meaningful dimension(s) that could not be reliably detected in the current sample. Finally, site-related factors, including differences in cohort composition, scanner platforms, and acquisition protocols, may have introduced residual variability despite harmonization using ComBat-GAM. Future studies using large single-site cohorts with standardized imaging protocols will be valuable for further confirming the robustness of the identified heterogeneity dimensions. This study characterized whole-brain neuroanatomy by jointly analyzing gray matter, white matter, and CSF/ventricular tissues. Future studies incorporating tau PET and amyloid PET biomarkers could further elucidate the structural and molecular mechanisms underlying each dimension, particularly the relative preservation of brain structure observed in Dimension 2 patients despite clinically meaningful impairment. Future studies should also evaluate the generalizability of these heterogeneity signatures through out-of-sample prediction in independent cohorts and more diverse populations. Finally, investigating whether dimensional membership and individual-level signature expression are associated with differential treatment response may be an important direction to inform precision medicine approaches and improve participant stratification in AD clinical trials.

## 5 CONCLUSIONS

Using a large multi-cohort sample and robust reproducibility approaches, we identified two stable neuroanatomical dimensions of AD – one marked by widespread atrophy and greater cognitive, clinical, functional, and genetic burden, and one with relatively preserved brain structure despite clinically meaningful impairment. Quantifying individual-level expression of these signatures, rather than relying on categorical assignment alone, captured clinically and genetically meaningful variation that a single diagnostic label obscures. These findings support the existence of biologically distinct AD dimensions and argue for incorporating neuroimaging-derived heterogeneity signatures into future patient stratification and precision-medicine approaches.

## Data Availability

All data and codes produced in the present study are available upon reasonable request to the authors.

## 6 ACKNOWLEDGEMENTS

Data collection and sharing for this project was funded by the Alzheimer’s Disease Neuroimaging Initiative (ADNI) (National Institutes of Health Grant U01 AG024904) and DOD ADNI (Department of Defense award number W81XWH-12-2-0012). ADNI is funded by the National Institute on Aging, the National Institute of Biomedical Imaging and Bioengineering, and through generous contributions from the following: AbbVie, Alzheimer’s Association; Alzheimer’s Drug Discovery Foundation; Araclon Biotech; BioClinica, Inc.; Biogen; Bristol-Myers Squibb Company; CereSpir, Inc.; Cogstate; Eisai Inc.; Elan Pharmaceuticals, Inc.; Eli Lilly and Company; EuroImmun; F. Hoffmann-La Roche Ltd and its affiliated company Genentech, Inc.; Fujirebio; GE Healthcare; IXICO Ltd.; Janssen Alzheimer Immunotherapy Research & Development, LLC.; Johnson & Johnson Pharmaceutical Research & Development LLC.; Lumosity; Lundbeck; Merck & Co., Inc.; Meso Scale Diagnostics, LLC.; NeuroRx Research; Neurotrack Technologies; Novartis Pharmaceuticals Corporation; Pfizer Inc.; Piramal Imaging; Servier; Takeda Pharmaceutical Company; and Transition Therapeutics. The Canadian Institutes of Health Research is providing funds to support ADNI clinical sites in Canada. Private sector contributions are facilitated by the Foundation for the National Institutes of Health (www.fnih.org). The grantee organization is the Northern California Institute for Research and Education, and the study is coordinated by the Alzheimer’s Therapeutic Research Institute at the University of Southern California. ADNI data are disseminated by the Laboratory for Neuro Imaging at the University of Southern California.

The resources provided by the Charles F. and Joanne Knight Alzheimer Disease Research Center for doi: 10.17632/sf7gxzdn63 were supported by the National Institutes of Health under award numbers P30AG066444, P01AG03991, P01AG026276, R01AG078964, RF1AG044546, RF1AG071706, RF1AG058501, and P30NS048056. We extend our sincere gratitude to the Knight ADRC research participants and their study partners for their invaluable time, contributions, and dedication to advancing this research. We also acknowledge the Knight ADRC Cores and Projects for their essential efforts in conducting, curating, and disseminating these datasets.

## CODE AND DATA AVAILABILITY

The codes are available at https://github.com/ganchand/ADDimensions. Original HYDRA codes are available at https://github.com/evarol/HYDRA. Datasets are available upon reasonable request following applicable guidelines of human subjects’ protection.

