## Supplemental Document for "Machine learning-derived Alzheimer’s disease dimensions: neuroanatomical, cognitive, clinical, functional, and genetic risk heterogeneity signatures"

### Table of Contents


*A. ADNI Dataset Inclusion and Exclusion Criteria* ______ *3*
*B. Knight ADRC Dataset Inclusion and Exclusion Criteria* ______ *3*
*C. Image Preprocessing* ______ *5*
*D. Study Variables* ______ *6*
*E. Polygenic Risk Scores* ______ *7*
*F. HYDRA Training Procedure* ______ *8*
*G. Reproducibility Experiment Designs* ______ *9*
*H. CDR-SB Sensitivity Analysis* ______ *10*


### List of Tables


*Supplementary Table S1. Missing patient variables by cohort.* ______ *12*
*Supplementary Table S2. Sex-stratified comparison of clinical, cognitive, and laboratory variables within each AD dimension.* ______ *13*
*Supplementary Table S3. Cardiometabolic and cardiovascular risk factor profiles between AD dimensions.* ______ *15*
*Supplementary Table S4. PRS profiles comparing AD Dimension 1 and Dimension 2.* ______ *16*
*Supplementary Table S5. Correlation of individual AD signature expressions with patient continuous variables.* ______ *17*


### List of Figures


*Supplementary Figure S1. FDR-corrected neuroanatomical differences between CN and Dimension 1.* ______ *18*
*Supplementary Figure S2. Sex-stratified neuroanatomical differences between CN and each patient dimension.* ______ *19*
*Supplementary Figure S3. Regional neuroanatomical patterns of AD Dimensions 1 and 2 relative to CN in CDR-SB cleaned subset* ______ *20*

**List of Abbreviations**

| | |
|---|---|
| AD | Alzheimer’s disease |
| ADNI | The Alzheimer's Disease Neuroimaging Initiative |
| ADRC | The Charles F. and Joanne Knight Alzheimer Disease Research Center |
| APOE | apolipoprotein E |
| ARI | Adjusted Rand Index |
| BMI | Body Mass Index |
| CDR | Clinical Dementia Rating |
| CDR-SB | Clinical Dementia Rating - Sum of Boxes |
| CN | cognitively normal |
| EMCI | early mild cognitive impairment |
| FAQ | Functional Activities Questionnaire |
| GWAS | Genome-wide association studies |
| HYDRA | Heterogeneity through Discriminative Analysis |
| LMCI | late mild cognitive impairment |
| MCI | mild cognitive impairment |
| MMSE | Mini-mental State Examination |
| MNI | Montreal Neurological Institute |
| MRI | magnetic resonance imaging |
| PET | positron emission tomography |
| PRS | polygenic risk score |
| PT | patients |
| ROI | region of interest |
| SVM | support vector machine |

### A. ADNI Dataset Inclusion and Exclusion Criteria

The first dataset used in this work was obtained from the ADNI database (adni.loni.usc.edu). ADNI was launched in 2003 as a public-private partnership. The primary goal of ADNI has been to test whether serial MRI, along with PET, other biological markers, and clinical and neuropsychological assessment, can be combined to measure the progression of MCI or early AD. The selected ADNI dataset consists of 667 participants (308 females; age: 55.1–91.5 years old, mean = 72.83 years old), where 187 are CN, 6 are MCI, 172 are EMCI, 152 are LMCI, and 150 dementia participants due to AD.

ADNI enrolled participants aged 55–90 years who had a reliable study partner, were English- or Spanish-speaking, and were willing to undergo neuroimaging, lumbar puncture, and longitudinal follow-up, and had Hachinski ≤ 4. ADNI inclusion and exclusion criteria are described in detail at https://adni.loni.usc.edu/.

### B. Knight ADRC Dataset Inclusion and Exclusion Criteria

The second dataset used in the present study comes from the Knight ADRC at Washington University in St. Louis. This is one of the longest-running and largest NIH-funded Alzheimer's Disease Research Centers in the United States. Established in 1985, the Knight ADRC maintains a longitudinal registry of community-dwelling volunteers spanning CN older adults, individuals with mild cognitive impairment, and patients with symptomatic AD dementia. The cohort is characterized by deep longitudinal phenotyping and a high proportion of participants followed to autopsy, enabling neuropathological confirmation of diagnosis. In the present study, we utilized cross-sectional T1-weighted MRI data from 1,090 Knight ADRC participants, comprising CN and individuals with AD dementia. In this selected population, 606 are female (ages: 45.2–91.4 years, mean = 70.37 years). Of these participants, 921 are CN while the remaining 169 are AD participants at various cognitive decline stages grouped into one diagnosis group (AD).

The Knight ADRC data comprises two dementia categories: AD and non-AD dementia. In this study, participants categorized as AD dementia were included, while those with non-AD dementia were excluded. Informed consent was obtained from each subject in accordance with the local institutional guidelines for human participants. Participants were classified as either CN or having

AD dementia using a combination of CDR and clinician assessment of etiology. A global CDR score is calculated from individual box scores in each domain, based on collateral sources and participant interviews. A global CDR of 0 signifies the absence of dementia, while a CDR of 0.5, 1, 2, and 3 indicates very mild, mild, moderate, and severe dementia, respectively. CN participants had a CDR of 0, whereas participants with AD had a CDR of 0.5 or higher. At baseline, participants with a primary cause of dementia other than AD (e.g., vascular dementia, Lewy body dementia, frontotemporal dementia), Parkinson's disease, or comorbid disorders that precluded longitudinal participation were excluded.

In both cohorts, a small number of participants had baseline CDR-SB scores inconsistent with their diagnostic category (e.g., CN participants with CDR-SB = 0.5). This likely reflects score collection at a different time point than diagnosis assignment. The sensitivity analysis was performed excluding those participants to ensure the results are not confounded by such discrepancies.

A summary of ADNI-2 and Knight ADRC inclusion and exclusion criteria is described in the following table.

| | ADNI | Knight ADRC |
|---|---|---|
| CN | Inclusions: MMSE 24–30; CDR = 0 (Memory Box = 0); free of memory complaints; normal Logical Memory II scores (education-adjusted); no significant cognitive or functional impairment.<br><br>Exclusions: significant neurologic disease (Parkinson's, multi-infarct dementia, Huntington's, seizure disorder, etc.); or history of significant head trauma; or known structural brain abnormalities. | Inclusions: Global CDR = 0; no cognitive impairment by clinician assessment.<br><br>Exclusions: primary dementia etiology other than AD (vascular, Lewy body, frontotemporal); Parkinson's disease; comorbid disorders precluding longitudinal participation. |
| MCI | Inclusions: MMSE 24–30; CDR = 0.5 (Memory Box ≥ 0.5); subjective memory concern reported by subject, informant, or clinician; abnormal education-adjusted Logical Memory II scores; general cognition and | N/A |

| | | |
|---|---|---|
| | functional performance sufficiently preserved to preclude AD diagnosis.<br><br><u>Exclusions</u>: significant neurologic disease except suspected incipient AD; or history of significant head trauma; or known structural brain abnormalities. | |
| AD | <u>Inclusions</u>: MMSE 20–26; CDR 0.5 or 1.0; meets NINCDS/ADRDA criteria for probable AD.<br><u>Exclusions</u>: neurologic disease other than suspected AD; major psychiatric disorder; MRI contraindications. | <u>Inclusions</u>: CDR ≥ 0.5 with clinician-confirmed AD as primary etiology; global CDR 0.5–3 (very mild to severe).<br><br><u>Exclusions</u>: primary etiology other than AD (vascular, Lewy body, frontotemporal dementia); Parkinson's disease; autosomal dominant AD (referred to DIAN); comorbid disorders precluding participation. |

### C. Image Preprocessing

T1-weighted structural MR images underwent a standardized preprocessing pipeline consisting of skull stripping, N4 bias field correction, and reorientation to MNI space. Skull stripping removes non-brain tissue, including the skull and extracranial fat, that would otherwise confound volumetric measurements of the brain. N4 bias field correction addresses low-frequency intensity non-uniformities arising from magnetic field inhomogeneities, which, if uncorrected, can systematically distort regional volume estimates and degrade segmentation accuracy. Reorientation to a standard MNI coordinate space ensures consistent anatomical alignment across participants and acquisition sites, enabling valid cross-subject comparisons of regional brain volumes.

### D. Study Variables

#### a) Structural MRI brain volumes

The primary features used for clustering were the 145 MUSE-derived regional brain volumes described above. For each participant, ROI volumes were extracted following ComBat-GAM harmonization to remove site, scanner, sex, age, and intracranial volume effects. Age at MRI acquisition, biological sex, years of education, self-reported race, and ethnicity were recorded.

#### b) Clinical and functional scores

Global cognitive status was assessed using the MMSE and the CDR scale, including the global CDR score and CDR-SB. Global CDR yields a categorical severity rating (0, 0.5, 1, 2, 3), and CDR-SB yields a continuous composite ranging from 0 to 18 that aggregates scores across six semi-structured interview domains. Three domains assess cognitive function – memory, orientation, and judgment and problem-solving – and three assess functional independence, including community affairs, home and hobbies, and personal care. Each domain is scored from 0 (no impairment) to 3 (severe impairment). Higher CDR-SB scores indicate greater overall disease severity. Functional impairment was assessed using the FAQ. It is a 10-item tool used to measure an individual's ability to perform complex daily tasks.

#### *c) APOE*

*APOE* genotype was determined for all participants and encoded as *ε4* carrier status: non-carrier, heterozygous carrier, or homozygous carrier. PRS AD were computed at five GWAS p-value inclusion thresholds ($5\times10^{-8}$, $5\times10^{-5}$, 0.05, 0.5, and 1.0), providing a gradient from genome-wide significant to genome-wide inclusive genetic burden estimates.

#### d) Vascular and metabolic risk factors

Vascular risk factors were ascertained from structured medical history interviews and included hypertension in addition to cohort-specific variables. For Knight ADRC participants, these variables consist of atrial fibrillation, angina pectoris, congestive heart failure, myocardial infarction, hypercholesterolemia, and diabetes mellitus. For ADNI participants, medical history variables included history of stroke, history of cardiovascular disease, endocrine-metabolic conditions, and tobacco smoking. BMI was calculated from measured height and weight.

#### e) Blood laboratory values

For ADNI participants, fasting blood laboratory values were obtained from the ADNI biomarker repository, including serum glucose, triglycerides, total cholesterol, creatinine, and serum uric acid. These markers were included to characterize the metabolic and renal profiles associated with each neuroanatomical dimension.

### E. Polygenic Risk Scores

PRS aggregate the cumulative effect of risk alleles across the genome into a single continuous estimate of an individual's genetic liability for a given trait, with each allele weighted by its effect size from the most comprehensive available GWAS. Compared to evaluating individual variants in isolation, PRS captures a greater proportion of trait variance and provides a more precise characterization of individual risk. Beyond disease prognosis, PRS can inform early intervention strategies, guide clinical trial enrollment, and quantify the degree of genetic overlap between comorbid complex traits through shared pleiotropic effects.[1–3]

PRS for AD[4] were computed using PRSice v2.3, which implements a clumping-and-thresholding framework. Variants in linkage disequilibrium are pruned and only those surpassing a specified association p-value threshold are retained, effectively shrinking the contribution of sub-threshold variants to zero while leaving included effect sizes unmodified. Scores were computed at five p-value thresholds ($5\times10^{-8}$, $5\times10^{-5}$, 0.05, 0.5, and 1). Two versions of the PRS were generated: one inclusive of the *APOE* region and one with it excluded (GRCh38: 19:43907927 to 19:45908810), to distinguish *APOE*-dependent from *APOE*-independent genetic risk. All scores were standardized to zero mean and unit variance within the analytic sample. PRS calculation is as follows:

$$PRS_j = \sum_i S_i \times G_{ij} - \frac{mean(PRS)}{SD(PRS)}$$

where $S$ is the summary statistic of the $i^{\text{th}}$ effect allele and $G$ is the number of effect alleles observed for the $j^{\text{th}}$ individual.

Because the GWAS summary statistics used for PRS derivation were generated predominantly from European-ancestry cohorts[4], effect size estimates and the underlying linkage disequilibrium structure on which clumping is based may not generalize to other ancestries. The PRS characteristics of Dimension 1 and Dimension 2 for those whose race is classified as white are presented in **Supplementary Table S4**. Among European participants, AD Dimension 1 carried a significantly higher AD polygenic burden than AD Dimension 2.

### F. HYDRA Training Procedure

#### a) Algorithm Overview

HYDRA is a semi-supervised clustering algorithm that identifies patient dimensions by simultaneously learning k linear discriminative hyperplanes, each separating CN from a distinct patient dimension. Each patient is assigned to the dimension whose separating hyperplane yields the maximum signed margin distance, ensuring that every identified cluster is defined by what distinguishes it from normative brain structure rather than by arbitrary proximity in feature space.

Formally, HYDRA solves a multiple-instance SVM optimization problem: for a given $k$, it learns $k$ weight vectors $w_1, \dots, w_k$ and bias terms $b_1, \dots, b_k$ such that each hyperplane $w_j \cdot x + b_j = 0$ maximally separates CN participants from the patients assigned to cluster $j$. CN participants serve as a shared reference class that constrains the solution space and prevents degenerate solutions.

To explore signature expression, we computed the signed distance from that subject's feature vector $x_i$ to the $k$-th hyperplane for each subject $i$ and each dimension $k$ as:

$$s_{i,k} = w_k^{\top} x_i + b_k$$

This scalar, referred to as the signature expression score, captures the degree to which a subject's neuroanatomical profile aligns with the direction defined by dimension $k$: positive values indicate a profile shifted toward that dimension's characteristic pattern, while negative values indicate a profile shifted away.

Signature expression scores for each dimension were then standardized to zero mean and unit variance (z-scored) using the distribution of all participants, to allow comparison across dimensions and cohorts. The resulting z-scores, one per dimension per subject, were used in all subsequent correlation analyses with cognitive, functional, and genetic variables.

##### b) Input Features

The input to HYDRA consisted of the 145 harmonized regional brain volumes for each participant, with CN assigned a diagnostic label of −1 and all patient groups (MCI and AD dementia) assigned a label of +1. No covariate correction was applied within HYDRA itself, as site, sex, age, and intracranial volume effects had already been removed during the ComBat-GAM harmonization step.

##### c) Cross-Validation and Stability Assessment

To evaluate clustering reproducibility, HYDRA was run using 10-fold cross-validation repeated across 50 independent random partitions of the data. In each fold, HYDRA was trained on 90% of the data, and the learned hyperplanes were used to assign cluster labels to the held-out 10%. Clustering stability was quantified using ARIs, which measure the agreement between cluster label assignments across different folds of the same repetition, corrected for chance agreement. ARI ranges from 0 (no better than random) to 1 (perfect reproducibility). The mean ARI across all 50 repetitions was computed for each *k*, and the value of *k* yielding the highest mean ARI was selected as the optimal number of dimensions.

##### d) Hyperparameter Selection

The SVM regularization parameter *C* was set to 0.25, following the original HYDRA implementation and consistent with prior applications in neuroimaging-based subtyping studies. Smaller values of *C* yield sparser hyperplane solutions and greater regularization, reducing sensitivity to individual outlier participants. The range of *k* evaluated was 2 through 8. All analyses were implemented using the MATLAB HYDRA toolbox (https://github.com/evarol/HYDRA) executed on a high-performance computing server.

#### G. Reproducibility Experiment Designs

To rigorously validate that identified cluster solutions reflect genuine biological structure rather than algorithmic artifacts or spurious data patterns, we designed a series of simulation experiments representing four distinct experimental conditions:

**a) Condition 1 – Real data (full sample)**

HYDRA was applied to the complete dataset comprising all CN and all patient participants with their true ROI volume measurements. This condition represents the primary analysis and establishes the observed ARI distribution against which all other conditions are compared.

**b) Condition 2 – Split-sample validation**

The combined dataset was randomly partitioned into two independent halves of equal size, with HYDRA trained on the first half and cluster assignments predicted for the second held-out half.

**c) Condition 3 – Balanced real-data subsampling**

In each independent run, a random 50% subset of true patient participants was drawn without replacement and combined with an equal-sized random sample of CN, yielding a balanced 50:50 case-control dataset of fixed size.

**d) Condition 4 – Null condition with randomly labeled controls**

To establish a stringent empirical null distribution, all participants in each run were drawn exclusively from the CN pool. A random 50% of these participants were then relabeled as patients (fake PT), with no change to their brain volume measurements.

Statistical significance of the observed ARI at each $k$ was assessed by comparison to the null distribution derived from Condition 4. A $k$ value was considered to reflect non-random clustering only when its mean ARI significantly exceeded the null distribution.

### H. CDR-SB Sensitivity Analysis

The diagnostic label of CN is assigned based on clinical assessment at a given visit, yet CDR-SB scores are not always recorded at the same timepoint, raising the possibility that a subset of CN participants may have had subclinical cognitive changes at the time of imaging.

We found that the neuroanatomical profiles of the identified dimensions were robust to the removal of diagnostically ambiguous participants at the CN–patient boundary. After excluding 64 CN participants with non-zero baseline CDR-SB scores and 2 patient participants with a CDR-SB of zero, Cohen's d effect sizes derived from the cleaned subset were nearly identical to those from the full sample for both AD Dimension 1 ($\rho = 1.000$, $p < 0.001$) and AD Dimension 2 ($\rho = 0.995$, $p < 0.001$), see **Supplementary Figure S3**.

### Supplementary Table S1. Missing patient variables by cohort.

| | Combined | ADNI | Knight ADRC |
|---|---|---|---|
| *n* | 1757 | 667 | 1090 |
| CDR-SB | 38 (2.2%) | 0 (0.0%) | 38 (3.5%) |
| Global CDR | 146 (8.4%) | 108 (16.2%) | 38 (3.5%) |
| MMSE | 155 (8.9%) | 107 (16.0%) | 48 (4.4%) |
| FAQ[Δ] | 1199 (68.3%) | 109 (16.3%) | 1090 (100.0%) |
| BMI (kg/m²)[Δ] | 1091 (62.1%) | 1 (0.1%) | 1090 (100.0%) |
| RCT11 serum glucose[Δ] | 1116 (63.5%) | 26 (3.9%) | 1090 (100.0%) |
| RCT19 triglycerides (GPO)[Δ] | 1116 (63.5%) | 26 (3.9%) | 1090 (100.0%) |
| RCT20 cholesterol (high performance)[Δ] | 1116 (63.5%) | 26 (3.9%) | 1090 (100.0%) |
| RCT392 lab creatinine (rate blanked)[Δ] | 1116 (63.5%) | 26 (3.9%) | 1090 (100.0%) |
| RCT8 serum uric acid[Δ] | 1116 (63.5%) | 26 (3.9%) | 1090 (100.0%) |
| PRS AD | 41 (2.4%) | 9 (1.3%) | 32 (2.9%) |
| History of hypertension | 145 (8.3%) | 0 (0.0%) | 145 (13.3%) |
| History of stroke[Δ] | 1090 (62.1%) | 0 (0.0%) | 1090 (100.0%) |
| History of atrial fibrillation[†] | 812 (46.2%) | 667 (100.0%) | 145 (13.3%) |
| History of angina[†] | 812 (46.2%) | 667 (100.0%) | 145 (13.3%) |
| History of congestive heart failure[†] | 812 (46.2%) | 667 (100.0%) | 145 (13.3%) |
| History of hypercholesterolemia[†] | 812 (46.2%) | 667 (100.0%) | 145 (13.3%) |
| History of myocardial infarction[†] | 812 (46.2%) | 667 (100.0%) | 145 (13.3%) |
| History of smoking | 1090 (62.1%) | 0 (0.0%) | 1090 (100.0%) |
| History of cardiovascular diseases[Δ] | 1090 (62.1%) | 0 (0.0%) | 1090 (100.0%) |
| History of endocrine-metabolic[Δ] | 1090 (62.1%) | 0 (0.0%) | 1090 (100.0%) |
| Diabetes history[†] | 812 (46.2%) | 667 (100.0%) | 145 (13.3%) |
| *APOE ε4* allele frequency | 46 (2.7%) | 4 (0.6%) | 42 (3.9%) |

Values are presented as *n* (%). [†]Variables were available only in the Knight ADRC cohort. [Δ]Variables were available only in the ADNI cohort.

**Supplementary Table S2. Sex-stratified comparison of clinical, cognitive, and laboratory variables within each AD dimension.**

| | Dimension 1 | | | Dimension 2 | | |
|---|---|---|---|---|---|---|
| | Female | Male | Female versus Male | Female | Male | Female versus Male |
| *n* | 178 | 216 | | 107 | 149 | |
| Age, mean ± SD (*n*) | 71.69 ± 7.21 (178) | 74.04 ± 8.30 (216) | p = 0.002 ** | 72.01 ± 7.54 (107) | 73.34 ± 7.02 (149) | p = 0.099 ns |
| CDR-SB, mean ± SD (*n*) | 3.14 ± 1.99 (173) | 3.47 ± 2.11 (209) | p = 0.139 ns | 1.79 ± 1.44 (106) | 1.90 ± 1.37 (146) | p = 0.323 ns |
| Global CDR, mean ± SD (*n*) | 0.67 ± 0.35 (147) | 0.69 ± 0.30 (178) | p = 0.332 ns | 0.51 ± 0.24 (88) | 0.52 ± 0.23 (131) | p = 0.676 ns |
| CDR memory, mean ± SD (*n*) | 0.80 ± 0.38 (178) | 0.84 ± 0.39 (215) | p = 0.373 ns | 0.58 ± 0.28 (107) | 0.60 ± 0.20 (149) | p = 0.226 ns |
| CDR orientation, mean ± SD (*n*) | 0.59 ± 0.48 (178) | 0.63 ± 0.45 (215) | p = 0.339 ns | 0.28 ± 0.36 (107) | 0.31 ± 0.35 (149) | p = 0.490 ns |
| CDR judgment & problem solving, mean ± SD (*n*) | 0.62 ± 0.43 (178) | 0.69 ± 0.43 (215) | p = 0.129 ns | 0.38 ± 0.29 (107) | 0.42 ± 0.34 (149) | p = 0.444 ns |
| CDR community affairs, mean ± SD (*n*) | 0.47 ± 0.47 (178) | 0.54 ± 0.50 (215) | p = 0.206 ns | 0.17 ± 0.31 (107) | 0.23 ± 0.29 (149) | p = 0.041 * |
| CDR home & hobbies, mean ± SD (*n*) | 0.51 ± 0.51 (178) | 0.58 ± 0.44 (215) | p = 0.038 * | 0.29 ± 0.40 (107) | 0.26 ± 0.31 (149) | p = 0.965 ns |
| CDR personal care, mean ± SD (*n*) | 0.08 ± 0.28 (178) | 0.22 ± 0.44 (215) | p < 0.001 | 0.05 ± 0.21 (107) | 0.05 ± 0.23 (149) | p = 0.805 ns |
| MMSE, mean ± SD (*n*) | 24.79 ± 3.95 (145) | 24.63 ± 3.64 (177) | p = 0.443 ns | 27.62 ± 2.56 (88) | 27.02 ± 3.00 (131) | p = 0.090 ns |
| FAQ$^{\Delta}$, mean ± SD (*n*) | 8.75 ± 8.43 (102) | 9.50 ± 7.52 (114) | p = 0.281 ns | 2.37 ± 4.46 (67) | 3.95 ± 5.23 (106) | p = 0.014 * |
| BMI (kg/m²)$^{\Delta}$, mean ± SD (*n*) | 26.25 ± 5.98 (128) | 27.07 ± 4.43 (146) | p = 0.011 ns | 27.56 ± 6.45 (85) | 27.64 ± 4.00 (121) | p = 0.922 ns |
| Years of education, mean ± SD (*n*) | 15.37 ± 2.63 (178) | 16.14 ± 2.83 (215) | p = 0.003 ** | 15.39 ± 2.86 (107) | 16.52 ± 2.66 (149) | p = 0.003 ** |
| RCT11 serum glucose$^{\Delta}$, mean ± SD (*n*) | 97.48 ± 23.59 (119) | 95.48 ± 31.76 (137) | p = 0.400 ns | 95.56 ± 21.58 (81) | 100.63 ± 29.03 (118) | p = 0.331 ns |
| RCT19 triglycerides (GPO)$^{\Delta}$, mean ± SD (*n*) | 129.05 ± 69.80 (119) | 124.35 ± 74.09 (137) | p = 0.661 ns | 134.90 ± 84.59 (81) | 147.36 ± 77.13 (118) | p = 0.167 ns |
| RCT20 cholesterol (high performance)$^{\Delta}$, mean ± SD (*n*) | 209.28 ± 43.63 (119) | 171.37 ± 51.49 (137) | p < 0.001 | 205.01 ± 50.21 (81) | 180.14 ± 37.41 (118) | p < 0.001 |
| RCT392 lab creatinine (rate blanked)$^{\Delta}$, mean ± SD (*n*) | 0.88 ± 0.24 (119) | 0.97 ± 0.49 (137) | p < 0.001 | 0.84 ± 0.34 (81) | 1.04 ± 0.29 (118) | p < 0.001 |
| RCT8 serum uric acid$^{\Delta}$, mean ± SD (*n*) | 4.65 ± 1.31 (119) | 5.36 ± 1.89 (137) | p < 0.001 | 4.64 ± 1.51 (81) | 5.73 ± 1.32 (118) | p < 0.001 |
| PRS AD including *APOE*, mean ± SD (*n*), mean ± SD (*n*) | 0.79 ± 1.07 (176) | 0.54 ± 1.16 (209) | p = 0.016 * | 0.17 ± 0.96 (105) | 0.29 ± 1.09 (147) | p = 0.445 ns |
| PRS $p < 5\times10^{-5}$, including *APOE*, mean ± SD (*n*) | 0.61 ± 0.94 (176) | 0.45 ± 1.10 (209) | p = 0.117 ns | 0.25 ± 1.01 (105) | 0.24 ± 1.06 (147) | p = 0.930 ns |
| PRS p < 0.05, including *APOE*, mean ± SD (*n*) | 0.02 ± 0.79 (176) | -0.03 ± 0.81 (209) | p = 0.448 ns | 0.21 ± 1.20 (105) | 0.00 ± 0.76 (147) | p = 0.554 ns |
| PRS p < 0.5, including *APOE*, mean ± SD (*n*) | 0.09 ± 0.89 (176) | 0.03 ± 0.85 (209) | p = 0.428 ns | 0.15 ± 1.09 (105) | 0.06 ± 0.77 (147) | p = 0.954 ns |

| | | | | | | |
|---|---|---|---|---|---|---|
| PRS $p < 1$, including *APOE*, mean ± SD (*n*) | 0.09 ± 0.89 (176) | 0.03 ± 0.86 (209) | p = 0.323 ns | 0.15 ± 1.09 (105) | 0.05 ± 0.76 (147) | p = 0.925 ns |
| PRS AD excluding *APOE*, mean ± SD (*n*) | 0.24 ± 1.02 (176) | 0.26 ± 1.08 (209) | p = 0.882 ns | 0.11 ± 0.96 (105) | 0.09 ± 1.09 (147) | p = 0.879 ns |
| PRS $p < 5\times10^{-5}$, excluding *APOE*, mean ± SD (*n*) | 0.05 ± 0.85 (176) | 0.10 ± 0.95 (209) | p = 0.792 ns | 0.20 ± 1.01 (105) | 0.00 ± 0.99 (147) | p = 0.185 ns |
| PRS $p < 0.05$, excluding *APOE*, mean ± SD (*n*) | -0.02 ± 0.79 (176) | -0.06 ± 0.81 (209) | p = 0.657 ns | 0.20 ± 1.19 (105) | -0.01 ± 0.76 (147) | p = 0.415 ns |
| PRS $p < 0.5$, excluding *APOE*, mean ± SD (*n*) | 0.06 ± 0.89 (176) | 0.01 ± 0.86 (209) | p = 0.545 ns | 0.14 ± 1.08 (105) | 0.05 ± 0.76 (147) | p = 0.999 ns |
| PRS $p < 1$, excluding *APOE*, mean ± SD (*n*) | 0.06 ± 0.89 (176) | 0.01 ± 0.87 (209) | p = 0.422 ns | 0.14 ± 1.08 (105) | 0.04 ± 0.76 (147) | p = 0.898 ns |
| History of hypertension, *n* (%) | 78 (45.1%) | 105 (53.0%) | p = 0.155 ns | 49 (47.1%) | 73 (49.7%) | p = 0.788 ns |
| History of stroke$^{\Delta}$, *n* (%) | 0 (0.0%) | 0 (0.0%) | N/A | 0 (0.0%) | 0 (0.0%) | N/A |
| History of atrial fibrillation$^{\dagger}$, *n* (%) | 1 (2.2%) | 1 (1.9%) | p = 1.000 ns | 2 (10.5%) | 3 (11.5%) | p = 1.000 ns |
| History of angina$^{\dagger}$, *n* (%) | 0 (0.0%) | 1 (1.9%) | p = 1.000 ns | 0 (0.0%) | 0 (0.0%) | N/A |
| History of congestive heart failure$^{\dagger}$, *n* (%) | 0 (0.0%) | 0 (0.0%) | N/A | 0 (0.0%) | 1 (3.8%) | p = 1.000 ns |
| History of hypercholesterolemia$^{\dagger}$, *n* (%) | 23 (51.1%) | 36 (69.2%) | p = 0.081 ns | 12 (63.2%) | 16 (61.5%) | p = 1.000 ns |
| History of myocardial infarction$^{\dagger}$, *n* (%) | 0 (0.0%) | 3 (5.8%) | p = 0.294 ns | 0 (0.0%) | 3 (11.5%) | p = 0.354 ns |
| History of smoking, *n* (%) | 45 (35.2%) | 69 (47.3%) | p = 0.057 ns | 31 (36.5%) | 41 (33.9%) | p = 0.814 ns |
| History of cardiovascular diseases$^{\Delta}$, *n* (%) | 77 (60.2%) | 100 (68.5%) | p = 0.189 ns | 51 (60.0%) | 85 (70.2%) | p = 0.168 ns |
| History of endocrine-metabolic$^{\Delta}$, *n* (%) | 67 (52.3%) | 57 (39.0%) | p = 0.037 * | 43 (50.6%) | 54 (44.6%) | p = 0.483 ns |

$^{\dagger}$Variables were available only in the Knight ADRC cohort. $^{\Delta}$Variables were available only in the ADNI cohort. ns, not significant; * $p < 0.05$; ** $p < 0.01$.

**Supplementary Table S3. Cardiometabolic and cardiovascular risk factor profiles between AD dimensions.**

| | Dimension 1 | Dimension 2 | Dimension 1 versus Dimension 2 |
|---|---|---|---|
| RCT11 serum glucose$^{\Delta}$, mean ± SD | 96.41 ± 28.28 | 98.56 ± 26.37 | U = 24474, p = 1.000 ns |
| RCT19 triglycerides (GPO)$^{\Delta}$, mean ± SD | 126.54 ± 72.16 | 142.29 ± 80.48 | U = 22410, p = 0.083 ns |
| RCT20 cholesterol (high performance)$^{\Delta}$, mean ± SD | 188.99 ± 51.59 | 190.26 ± 44.78 | U = 25586, p = 1.000 ns |
| RCT392 creatinine (Rate Blanked)$^{\Delta}$, mean ± SD | 0.93 ± 0.39 | 0.96 ± 0.32 | U = 25286, p = 1.000 ns |
| RCT8 serum uric acid$^{\Delta}$, mean ± SD | 5.03 ± 1.68 | 5.29 ± 1.50 | U = 23370, p = 0.392 ns |
| History of hypertension, *n* (%) | 183 (49.3%) | 122 (48.6%) | $\chi^2$ = 0.009, df = 1, p = 1.000 ns |
| History of stroke$^{\Delta}$, *n* (%) | 0 (0.0%) | 0 (0.0%) | $\chi^2$ = 0.000, df = 1, p = 1.000 ns |
| History of atrial fibrillation$^{\dagger}$, *n* (%) | 2 (2.1%) | 5 (11.1%) | $\chi^2$ = 3.614, df = 1, p = 0.172 ns |
| History of angina$^{\dagger}$, *n* (%) | 1 (1.0%) | 0 (0.0%) | $\chi^2$ = 0.000, df = 1, p = 1.000 ns |
| History of congestive heart failure$^{\dagger}$, *n* (%) | 0 (0.0%) | 1 (2.2%) | $\chi^2$ = 0.156, df = 1, p = 1.000 ns |
| History of hypercholesterolemia$^{\dagger}$, *n* (%) | 59 (60.8%) | 28 (62.2%) | $\chi^2$ = 0.475, df = 2, p = 1.000 ns |
| History of myocardial infarction$^{\dagger}$, *n* (%) | 3 (3.1%) | 3 (6.7%) | $\chi^2$ = 0.288, df = 1, p = 1.000 ns |
| History of smoking$^{\Delta}$, *n* (%) | 114 (41.6%) | 72 (35.0%) | $\chi^2$ = 1.922, df = 1, p = 0.497 ns |
| History of cardiovascular diseases$^{\Delta}$, *n* (%) | 177 (64.6%) | 136 (66.0%) | $\chi^2$ = 0.051, df = 1, p = 1.000 ns |
| History of endocrine-metabolic$^{\Delta}$, *n* (%) | 124 (45.3%) | 97 (47.1%) | $\chi^2$ = 0.094, df = 1, p = 1.000 ns |
| Diabetes history$^{\dagger}$ | | | $\chi^2$ = 0.8948, df = 1, p = 0.344 ns |
| · Absent, *n* (%) | 89 (91.8%) | 39 (86.7%) | |
| · Remote/inactive, *n* (%) | 8 (8.2%) | 6 (13.3%) | |

$^{\dagger}$Variables were available only in the Knight ADRC cohort. $^{\Delta}$Variables were available only in the ADNI cohort. ns, not significant.

**Supplementary Table S4. PRS profiles comparing AD Dimension 1 and Dimension 2.**

| | Dimension 1 | Dimension 2 | Dimension 1 versus Dimension 2 |
|---|---|---|---|
| | *All races (n = 637)* | | |
| PRS $p < 5\times10^{-5}$, including *APOE* | 0.52 ± 1.03 | 0.24 ± 1.04 | U = 56814, $p < 0.001$ |
| PRS $p < 0.05$, including *APOE* | -0.01 ± 0.80 | 0.09 ± 0.97 | U = 46705, p = 1.000 ns |
| PRS $p < 0.5$, including *APOE* | 0.05 ± 0.87 | 0.10 ± 0.91 | U = 48289, p = 1.000 ns |
| PRS $p < 1$, including *APOE* | 0.06 ± 0.88 | 0.09 ± 0.91 | U = 48432, p = 1.000 ns |
| PRS $p < 5\times10^{-5}$, excluding *APOE* | 0.08 ± 0.91 | 0.09 ± 1.00 | U = 49208, p = 1.000 ns |
| PRS $p < 0.05$, excluding *APOE* | -0.04 ± 0.80 | 0.08 ± 0.97 | U = 45302, p = 0.473 ns |
| PRS $p < 0.5$, excluding *APOE* | 0.03 ± 0.87 | 0.09 ± 0.91 | U = 47554, p = 1.000 ns |
| PRS $p < 1$, excluding *APOE* | 0.03 ± 0.88 | 0.08 ± 0.91 | U = 47801, p = 1.000 ns |
| | *European participants (n = 590)* | | |
| PRS $p < 5\times10^{-8}$, including *APOE* | 0.67 ± 1.14 | 0.24 ± 1.07 | U = 50594, $p < 0.001$ |
| PRS $p < 5\times10^{-8}$, excluding *APOE* | 0.27 ± 1.07 | 0.13 ± 1.05 | U = 44215, p = 0.117 ns |

Values are presented as mean ± standard deviation. *APOE*-inclusive and *APOE*-exclusive PRS were calculated using the indicated GWAS significance thresholds. ns, not significant.

**Supplementary Table S5. Correlation of individual AD signature expressions with patient continuous variables.**

| | AD Signature | Spearman's ρ | p-value | |
|---|---|---|---|---|
| CDR-SB | 1 | 0.447 | < 0.001 | |
| | 2 | -0.110 | 0.006 | ** |
| Global CDR | 1 | 0.358 | < 0.001 | |
| | 2 | -0.121 | 0.005 | ** |
| CDR Memory | 1 | 0.421 | <0.001 | |
| | 2 | -0.102 | 0.009 | ** |
| CDR Orientation | 1 | 0.412 | <0.001 | |
| | 2 | -0.074 | 0.060 | ns |
| CDR Judgment & Problem Solving | 1 | 0.404 | <0.001 | |
| | 2 | -0.103 | 0.009 | ** |
| CDR Community Affairs | 1 | 0.395 | <0.001 | |
| | 2 | -0.108 | 0.006 | ** |
| CDR Home & Hobbies | 1 | 0.343 | < 0.001 | |
| | 2 | -0.102 | 0.009 | ** |
| CDR Personal Care | 1 | 0.209 | < 0.001 | |
| | 2 | -0.074 | 0.058 | ns |
| BMI ($kg/m^2$)$^{\Delta}$ | 1 | -0.136 | 0.003 | ** |
| | 2 | 0.005 | 0.916 | ns |
| Years of Education | 1 | -0.080 | 0.041 | * |
| | 2 | 0.079 | 0.044 | * |
| RCT11 Lab Serum Glucose$^{\Delta}$ | 1 | -0.059 | 0.213 | ns |
| | 2 | 0.020 | 0.670 | ns |
| RCT19 Lab Triglycerides (GPO)$^{\Delta}$ | 1 | -0.119 | 0.011 | * |
| | 2 | 0.022 | 0.639 | ns |
| RCT20 Lab Cholesterol (High Performance)$^{\Delta}$ | 1 | -0.003 | 0.942 | ns |
| | 2 | 0.034 | 0.473 | ns |
| RCT392 Lab Creatinine (Rate Blanked)$^{\Delta}$ | 1 | -0.022 | 0.638 | ns |
| | 2 | -0.036 | 0.439 | ns |
| RCT8 Lab Serum Uric Acid$^{\Delta}$ | 1 | -0.103 | 0.028 | * |
| | 2 | 0.014 | 0.774 | ns |
| PRS AD excluding *APOE* | 1 | 0.075 | 0.058 | ns |
| | 2 | -0.014 | 0.725 | ns |

$^{\Delta}$Variables were available only in the ADNI cohort. ns, not significant; * $p < 0.05$; ** $p < 0.01$.

**Supplementary Figure S1. FDR-corrected neuroanatomical differences between CN and Dimension 1.**

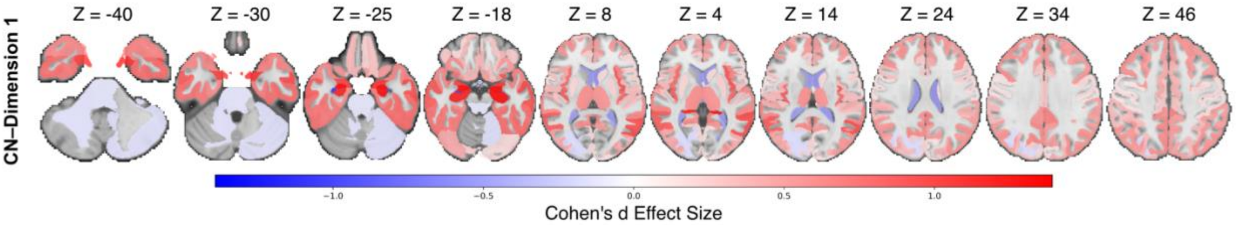


Cohen's d effect size maps comparing CN and AD Dimension 1 across 145 gray and white matter regions. Of 118 regions nominally significant at $p < 0.05$, 113 survived FDR correction ($q < 0.05$) and are displayed. The five regions that did not survive correction were the right occipital pole, left cerebellum white matter, right fornix, right triangular part of the inferior frontal gyrus, and cerebellar vermal lobules. Dimension 2 is not shown because no regions survived FDR correction. Positive values (red) indicate lower regional volumes in Dimension 1 relative to CN, whereas negative values (blue) indicate higher regional volumes. Slice positions are shown in MNI z-coordinates.

**Supplementary Figure S2. Sex-stratified neuroanatomical differences between CN and each patient dimension.**

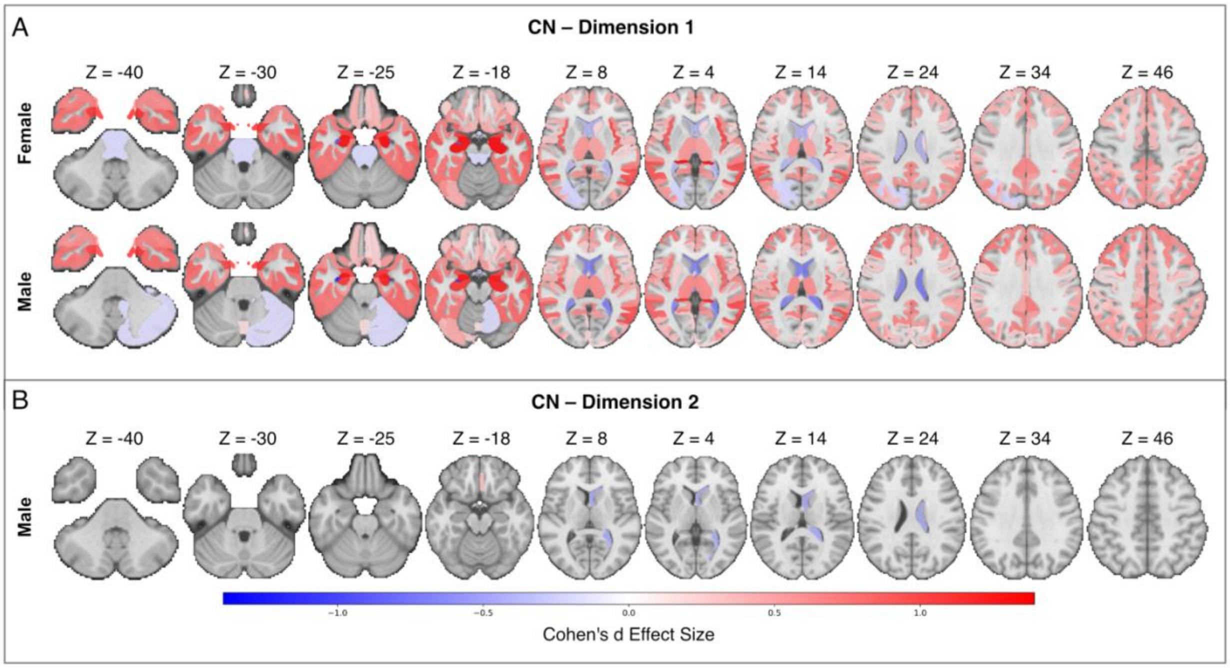


Cohen's d effect size maps comparing CN with AD Dimension 1 (A) and Dimension 2 (B), stratified by sex. Dimension 1 showed widespread volumetric reductions relative to CN in both sexes, with males exhibiting a greater number of significantly affected regions (108 vs. 91 FDR-corrected ROIs), spanning cortical, subcortical, and cerebellar areas. Dimension 2 displayed minimal structural differences, with only 2 regions surviving FDR correction in males, including the ventricles, and none in females. Positive values (red) indicate lower regional volumes relative to CN, whereas negative values (blue) indicate higher regional volumes. Slice positions are shown in MNI z-coordinates.

**Supplementary Figure S3. Regional neuroanatomical patterns of AD Dimensions 1 and 2 relative to CN in CDR-SB cleaned subset**

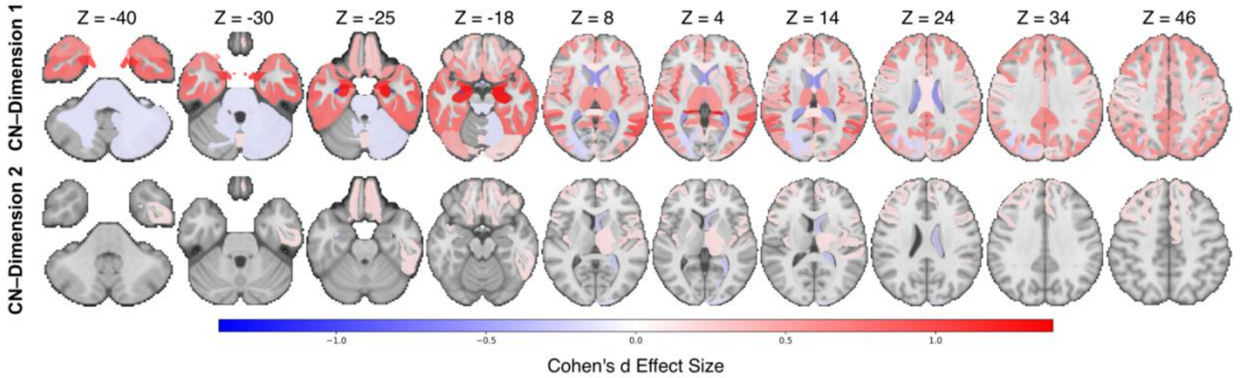


Regional Cohen's *d* effect size maps comparing Dimension 1 and Dimension 2 with CN (n = 1044), excluding 64 CN with non-zero CDR-SB and 2 patients with zero CDR-SB, across 145 brain regions. This subset exhibits largely similar neuroanatomical patterns compared to the full dataset. Positive values (red) indicate regional volume reduction relative to CN, whereas negative values (blue) indicate regional volume increase. Only regions with $p < 0.05$ are displayed, and color intensity represents Cohen's *d* effect size. Slice positions are shown in MNI *z*-coordinates.
